# Developing the PATH-GP (Prevention and Testing for HIV in General Practice) intervention: a Person-Based Approach intervention development study to increase HIV testing and PrEP access

**DOI:** 10.1101/2025.03.03.25323009

**Authors:** Anne Scott, Hannah Family, Jeremy Horwood, John Saunders, Ann Sullivan, Jo Burgin, Lindsey Harryman, Sarah Stockwell, Joanna Copping, Paul Sheehan, John MacLeod, Sarah Dawson, Joanna May Kesten, Sarah Denford

**Affiliations:** The National Institute for Health and Care Research Applied Research Collaboration West (NIHR ARC West) at University Hospitals Bristol and Weston NHS Foundation Trust, UK; Population Health Sciences, Bristol Medical School, University of Bristol, Bristol, UK; National Institute for Health and Care Research, Health Protection Research Unit (HPRU) in Behavioural Science and Evaluation. Population Health Sciences, Bristol Medical School, University of Bristol, Bristol, UK; Centre for Academic Primary Care (CAPC), Bristol Medical School, University of Bristol, Bristol, UK; Blood Safety, Hepatitis, STI and HIV Division, UK Health Security Agency, UK; Chelsea and Westminster Hospital NHS Foundation Trust, London, UK; Unity Sexual Health, University Hospitals Bristol and Weston NHS Foundation Trust, Bristol, UK; Communities and Public Health, Bristol City Council, Bristol, UK; Public Health and Prevention, Bath and North East Somerset Council, UK

**Keywords:** General Practice, Family practice, HIV Testing, Pre-Exposure Prophylaxis, Behavior-change Interventions, Qualitative Research

## Abstract

**Background:** Testing for HIV, linkage to treatment and access to pre-exposure prophylaxis (PrEP) (medication which reduces the risk of acquiring HIV) is essential for early HIV diagnosis, treatment, and prevention. General practice could play a key role in maximising H IV testing opportunities and supporting access to PrEP.

**Aim:** To develop an intervention for general practice to increase HIV testing and facilitate access to PrEP.

**Design and setting:** A person-based approach (PBA) intervention development study using the Capability, Opportunity, Motivation, Behaviour (COM-B) Model in South West England.

**Method:** A scoping review and semi-structured interviews with healthcare professionals (HCPs) and local organisation representatives with an interest in HIV prevention/healthcare) were conducted to understand the challenges and find potential solutions to increase HIV testing and facilitate access to PrEP in general practice. Intervention development used focus groups with HCPs and the public. Purposive sampling ensured diversity of practices and participants. Data was analysed using the PBA table of planning and CLIP-Q approach.

**Results:** Barriers identified included lack of clinician knowledge of HIV and PrEP, concern about stretched resources and a lack of systematic testing methods. Proposed strategies included simpler testing methods to normalise testing and reduce HIV stigma. The intervention developed consists of: education, a prompt to test, simplified and standardised testing and PrEP signposting processes, patient information, and practice champions.

**Conclusion:** Research is needed to explore the feasibility and the effectiveness of this multicomponent intervention to increase testing and access to PrEP within general practice. Funding barriers also need to be addressed.

**How this fits in:**

- General practice could play a key role in maximising HIV testing opportunities and supporting access to pre-exposure prophylaxis (PrEP). Opportunities to carry out HIV testing continue to be missed in general practice leading to late HIV diagnosis which is associated with reduced life expectancy, increased mortality and greater treatment costs.
- Patient acceptability for HIV testing is high but testing rates are low and variable and patients experience barriers to accessing PrEP through sexual health clinics.
- This research reports the development of a multi-faceted approach to increase HIV testing and access to PrEP in general practice using the person-based approach. This included investigating public and healthcare professional perceptions about a range of approaches including training, opt-out testing, and the provision of decision-making aids.
- Targeting capability, opportunity and motivation barriers the intervention encompasses HIV and PrEP education and training, and the provision of simpler and systematic approaches to testing.

## Introduction

The UNAIDS global goal to end new HIV infections by 2030 (1), requires coordinated efforts to improve HIV prevention, detection and treatment. In the UK, this is a high-priority for NHS England and the Department of Health and Social Care as outlined in the HIV Action Plan (2) for England and National Institute for Health and Care Excellence (NICE) guidance (3, 4).

In 2023 4,700 people in England were estimated to be living with undiagnosed HIV (5) and 923 of people first diagnosed in England were diagnosed late (5, 6). Reducing late HIV diagnosis (defined as a CD4 count of <350 cells within 91 days of diagnosis) and missed opportunities for HIV testing are central to the elimination of HIV transmission (2–7). Late HIV diagnosis is associated with increased risk of hospitalisation, reduced life expectancy, increased mortality and greater treatment and secondary care costs (8–10). Conversely, HIV anti-retroviral therapy (ART) increases life expectancy in line with the general population (11) and those on effective treatment with an undetectable viral load cannot pass HIV on (12). Routine HIV testing in high HIV prevalence areas is cost-effective in the medium term (13). Studies demonstrate high levels of patient acceptance and acceptability of HIV testing through general practice (14–17). General practice could therefore play a central role in early HIV diagnoses. Despite this, HIV testing rates in general practice are low and variable (18, 19) and patients often present with HIV related indicator conditions (ICs) several times before being diagnosed with HIV (20, 21) and primary or early-stage HIV infection is frequently missed (22).

In areas of high prevalence (>2 per 1000 people aged 15-59 (e.g. Bristol 2.37 per 1000 people) (23) and extremely high prevalence (>5 per 1000 people aged 15-59 guidance from the NICE and British HIV Association (BHIVA) (3, 24, 25) recommends offering testing to all new registrants, to patients presenting with ICs and to patients undergoing blood tests for any other reason. Offering testing in high HIV prevalence areas according to this guidance is cost-effective in the medium term (13). In addition, in areas of extremely high prevalence HIV testing should be considered opportunistically at each consultation. In low prevalence areas, guidelines recommend testing in response to ICs and testing people who may be exposed to HIV (25). HIV testing is important in in all areas (26) and requires adequate resources and commissioning(16) to pay for staff training and increased testing. Evidence suggests that testing outside specialist GUM/sexual health services (SHS) does not fully adhere to these guidelines (17, 27).

Increasing general practice HIV testing in line with recommendations above also provides opportunities to identify patients eligible for pre-exposure prophylaxis (PrEP) (28) a biomedical intervention that reduces the risk of getting HIV from sex by 99% (7, 28, 29). Increasing access to PrEP, is an important, potentially cost-effective (4, 30) means of preventing new HIV infections. PrEP is currently only available via NHS specialist level 3 GUM services which leads to inequalities in access amongst some patient groups which use SHS less (6) and calls have been made for interventions to raise awareness of PrEP in women, black and ethnic minority and transgender populations (31) who were under-represented in the PrEP Impact trial (32).

General practice could play a crucial role in increasing PrEP access by informing patients about PrEP, and referring them to specialist services (4, 33). There is currently a dearth of evidence on whether GP HCPs in England are supporting access to PrEP (34) and a lack of interventions to support increased awareness of PrEP and signposting to PrEP services in general practice.

Education interventions for HCPs can increase HCP awareness, confidence, and consideration of HIV testing but may not address opportunity and motivation barriers to testing (35) and impacts on testing vary (19, 36–39). Multi-component, complex interventions, may enhance the effects of educational interventions on HIV testing behaviours (40, 41).

To increase HIV testing in high and low prevalence areas there is a need to utilise a validated approach to intervention development, incorporating behaviour change theory and interest holder involvement to maximise acceptability and effectiveness (42).

This paper presents the development of a complex, theory-based general practice intervention to increase HIV testing and support access to PrEP.

## Method

The study, conducted between October 2022-August 2024, was guided by the person-based approach (PBA) (42) and the collaborative and intensive pragmatic qualitative (CLIP-Q) approach (Fig 1) (43). PBA aims to develop interventions tailored to the views, needs and experiences of the individuals who use them. Using an iterative approach to planning and optimising, the intervention aims to be relevant, engaging and easy to use (42). CLIP-Q is a team-based approach to responsive qualitative research that, like PBA, emphasises collaboration (with public and key interest holders) at all stages of the design, conduct and implementation of the study. The theoretical framework underpinning the development of the intervention is the COM-B (‘capability’, ‘opportunity’, ‘motivation’ and ‘behaviour’) model (44), which provides a framework to understand the barriers and facilitators to increasing HIV testing and supporting access to PrEP in general practice. COM-B posits that a behaviour (B), for example, HIV testing, is influenced by an individual having capabilities (C) (skills and knowledge), opportunity (social and physical) and motivation (M) (automatic emotion and reflective beliefs, intentions).

**Fig 1.**
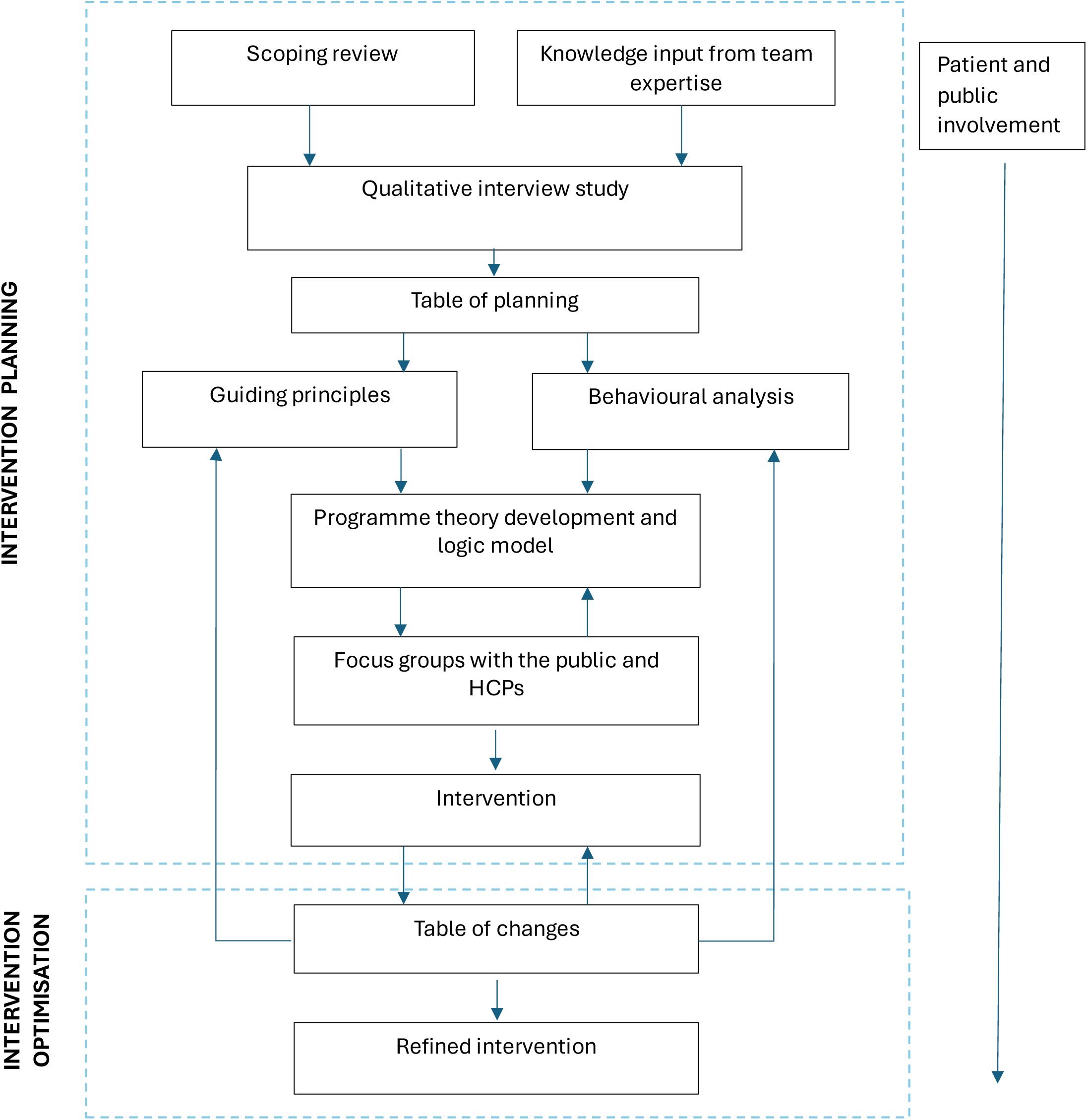
Person-based approach study design overview.

### Evidence and theory

A scoping review of systematic reviews to understand the barriers and facilitators to increasing HIV testing and access to PrEP in general practice was conducted. In accordance with the JBI (Joanna Briggs Institute) methodology for scoping reviews (45). Searches were performed in Ovid MEDLINE, Ovid APA PsycINFO, Web of Science Core Collection, Ovid Embase. Additionally, we searched the following review databases: Epistmonikos, Health Evidence, Database of Promoting Health Effectiveness Reviews (DoPHER) and NIHR Health Technology Assessments. All types of study were included (see example search strategies Supplementary Box S1). Databases were searched in December 2022 (PrEP search) and February 2023 (HIV testing search). A grey literature search was also conducted, including: Google, other relevant databases, charity, organisation, and healthcare websites.

### Patient and public involvement (PPI)

Three members of the public helped refine the study design and participant facing materials, topic guides, data analysis and intervention planning and optimisation.

Meetings were held with local interest holders involved in SHS and a charity supporting individuals living with HIV to support all aspects of the study.

### Qualitative Data Collection

#### Setting

General practices from low and high HIV prevalence areas in South West England (specifically, Bristol, North Somerset and South Gloucestershire; Gloucestershire; and Bath and North East Somerset).

#### Qualitative interviews with staff and interest holders

Semi-structured telephone or video interviews were conducted with GP HCPs (including GPs, practice pharmacists, practice nurses, phlebotomists) and interest holders (e.g. commissioners) to understand attitudes towards increasing HIV testing and access to PrEP in general practice.

#### Participants

The Clinical Research Network (CRN) invited all research active GP practices in the study areas. Those practices expressing an interest were selected across HIV testing data quartiles as a rate of practice population size, HIV prevalence (low vs high) using Summary Profile of Local Authority Sexual Health (SPLASH) reports (46), urban or rural area and indices of multiple deprivation.

#### Materials and Procedures

Twelve GP practices emailed their staff information about the study and interested staff contacted the research team to arrange an interview. The interview topic guide was informed by the results of the scoping review, and COM-B model (44) and was modified in response to issues raised in earlier interviews (see Supplementary Box S2). Interviews were audio recorded with consent, professionally transcribed and anonymised. Interview participants or their practices were reimbursed for their time according to the National Institute of Health and Care Research rates of pay.

#### Focus groups with staff and the public

Members of the public and HCPs were invited to separate online focus groups (of up to one hour duration) to refine the guiding principles for the intervention and co-develop the intervention content. Guiding principles present the key contextual issues and design objectives required to facilitate behaviour change and were developed iteratively.

Members of the public with a range of characteristics (e.g. gender, age, ethnicity) and experiences of HIV testing and/or PrEP use were recruited with the assistance of Health Ambassadors, public involvement and applied health researchers and their networks (with ability to reach diverse groups). This was to ensure we included the voices of those often underrepresented in research, those more likely to experience late HIV diagnosis and those who are less likely to attend sexual health clinics (e.g. people from ethnic minoritised communities).

GP HCPs with a mix of clinical backgrounds and experiences of HIV testing and/or PrEP delivery were recruited from those who took part in the interviews or expressed an interest but had not yet participated.

#### Analysis

Interviews were summarised immediately following recording using Microsoft® Excel® for Microsoft 365 (43) to chart knowledge of HIV and PrEP and barriers and facilitators to increasing HIV testing and PrEP access. This approach is particularly suitable when interviews are focused (43). Interview transcripts were used to supplement the initial analysis.

Key findings on barriers and facilitators extracted from the scoping review and interview findings were added to the PBA table of planning and mapped to the COM-B model of behaviour change (see Supplementary table S3) to identify approaches that would enhance individuals’ capability, create opportunities for behaviour change and provide motivation to increase HIV testing and access to PrEP. From this analysis a draft intervention, guiding principles for the intervention and an accompanying logic model were developed (see supplementary figure S4).

Feedback from the focus groups were documented in a PBA table of changes. Suggested changes were coded by importance in relation to the guiding principles, number of times the comment was raised and whether evidence from behaviour change theory suggests the change is likely to increase the chance of the behaviour being performed.

## Results

The scoping review to support access to PrEP, identified 339 publications of which 99 were excluded as duplicates, 163 were excluded based on title and abstract and 46 were excluded based on full text leaving 31 included.

The scoping review for HIV testing identified 393 publications of which 121 were excluded as duplicates, 205 were excluded based on title and abstract and a further 36 were excluded on full-text reading and a further 36 were excluded following full-text reading. 9 papers were included.

The scoping review and interview findings are presented in Supplementary Table S3.

25 HCPs (20 GPs, 4 practice nurses and one pharmacist) from 12 GP practices and 4 interest holders (representing SHS, an HIV charity, and commissioning organisations) participated in interviews lasting an average of 34 minutes. Tables 1 and 2 provide demographic information about GP practices and participants (to preserve anonymity interest holder demographics are not presented). Interview findings are framed using the COM-B model.

**Table 1.** Practice Characteristics.

| GP Practice | HIV testing quartile<sup>a</sup> | List Size <sup>b</sup> | IMD decile <sup>c</sup> | Area | HIV Prevalence | Staff |
| --- | --- | --- | --- | --- | --- | --- |
|  |  |  |  |  |  | Interviewed n |
| 1 | Quartile 4 | Medium | 4 | Sub-urban | High (2-<5) on the boundary with low (1-<2) | 4 |
| 2 | Quartile 2 | Medium | 10 | Sub-urban | Low (1-<2) | 1 |
| 3 | Quartile 4 | Medium | 5 | Sub-urban | Low (1-<2) on the boundary with high (2-<5) | 1 |
| 4 | Quartile 4 | Small | 10 | Sub-urban | Low (1<2) | 1 |
| 5 | Quartile 2 | Medium | 9 | Sub-urban | Low (1<2) in between high prevalence areas | 2 |
| 6 | Quartile 2 | Very Large | 5 | Urban | High (2-<5) | 2 |
| 7 | Quartile 1 | Very Large | 10 | Sub-urban and Rural | Low (<1) | 4 |
| 8 | Quartile 4 | Medium | 2 | Urban | High (2-<5) | 3 |
| 9 | Quartile 1 | Large | 8 | Urban | High (2-<5) | 1 |
| 10 | Quartile 1 | Large | 10 | Urban | Low (1-<2) | 3 |
| 11 | Quartile 4 | Small | 9 | Rural | Low <1 | 2 |
| 12 | Quartile 4 | Very Large | 5 | Urban | High (2-<5) | 1 |

<sup>a</sup>Quartile 1 = between minimum value and the lower quartile. Quartile 2 = between the lower quartile and the median, Quartile 3 = between the median and upper quartile, Quartile 4 = between the upper quartile and the maximum value. NB The values were tests per 1000 population for 2022.
<sup>b</sup>Small: < 10,000; Medium: 10,000 – 20,000; Large: 20,000-30,000; Very Large: >30,000
<sup>c</sup>1 = most deprived and 10 = most affluent.

**Table 2.**
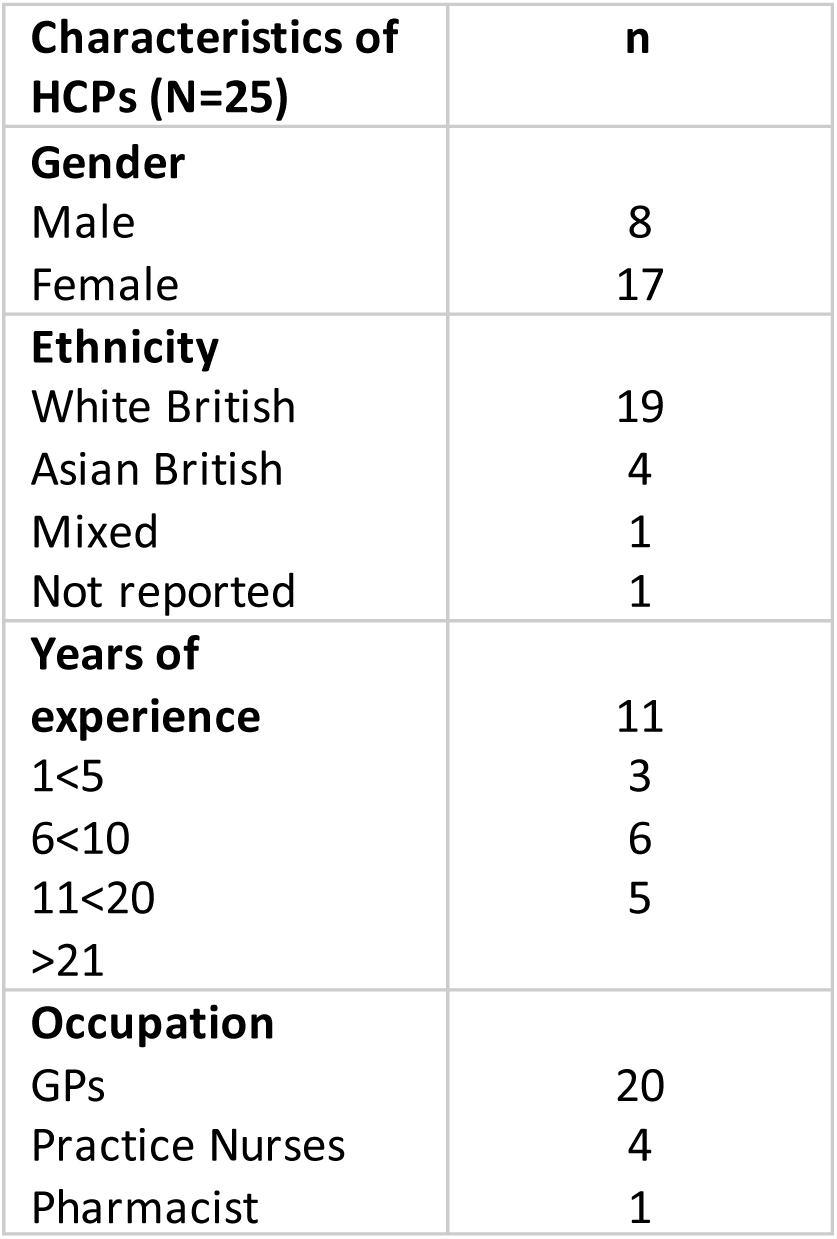
Demographic characteristics – HCP Interviews.

### Scoping review findings

Knowledge of HIV testing guidelines and prevalence in the practice population is low among HCPs (37, 47–49). HCPs also lack confidence and experience discomfort discussing HIV with patients and offering routine HIV tests (47, 48, 50, 51). Concerns about patient reactions and stigma act as a barrier to initiating conversations about HIV (48, 52). There was a lack of HCP awareness of the HIV stigma experienced by some minoritised groups (47).

HCPs view routine HIV testing as time consuming due to the belief that pre and post-test counselling and a lengthy consent process is required (47, 51, 53). HIV tests are not considered or included routinely when blood tests are requested (47) and HCPs do not (typically) have a systematic way of recording behaviours which increase the risk of exposure to HIV so may not always recognise opportunities for testing (48, 54).

HCPs may also not recognise or be familiar with ICs - and may not think about requesting an HIV test - particularly among those not considered typically ‘high risk’ (18, 47, 50, 55, 56).

Many of the PrEP reviews were conducted in the United States of America (USA) where the medication has been available longer than in England and where any HCP licensed to prescribe medication can prescribe PrEP. Despite this availability barriers to prescribing persist amongst primary care HCPs in the USA.

HCPs knowledge of PrEP is low (57–69). Most HCPs do not discuss PrEP with patients due to this lack of knowledge - as well as viewing PrEP as something that should be delivered by sexual health services (57, 59, 63, 64, 69–73).

USA primary care HCPs report a lack of skills, knowledge and time to counsel patients, to prescribe and carry out PrEP follow up (60, 61, 64, 68, 69).

HCPs experience discomfort discussing sexual activities/history with patients (57, 60, 62) and have concerns about PrEP use leading to risky behaviour (60, 61, 64, 68, 70, 74).

### Interview findings

#### Capability barriers

##### Knowledge and awareness

Most HCPs were unaware of the local prevalence of HIV and/or national testing guidelines with those in high prevalence area practices indicating that their practice did not carry out routine testing as per the guidelines. Participants reported that testing was carried out in response to indicator conditions e.g. unexplained weight loss, lymphadenopathy or risk factors indicating the need for an HIV test e.g. intravenous drug use.

HCPs felt that a lack of skills and confidence in having a conversation with patients about HIV specifically and sexual health in general was a barrier to testing for some staff although many did not feel they lacked these skills themselves.

A lack of knowledge and confidence among HCPs was also a barrier to discussing PrEP with patients. Most HCPs acknowledged that they knew very little about PrEP and few had discussed it with a patient.

HCPs expressed uncertainty about who might benefit from PrEP and felt that patients were better served by attending specialist services. Similarly, some participants felt that patients would know where to seek help.

> *I almost feel like people often know where to go for it and they know about it if they’re in a high-risk group.* (Participant 21 - GP - high prevalence area).

ICs were not known by all participants, and there was an acknowledgement that an inability to identify those who should be tested can lead to late HIV diagnosis.

> *GPs often can miss those potential symptoms as indicative of HIV infection. I think they always tend to be thinking, it’s probably got to be something else, and it might be the fifth or the sixth thing they consider, rather than the first or the second.* (Participant 26 - Interest Holder - low prevalence area)

Despite this, there was a reported lack of systematic approaches to identifying individuals at increased risk of HIV.

There was a lack of standardisation across practices regarding tests offered during sexually transmitted infection (STI) checks. Some HCPs understood the need to include HIV/blood borne virus (BBV) tests, but many preferred to refer patients to SHS.

> *If they were requesting a routine STI screen, we would do the swabs here, but for blood borne viruses, we would generally refer to (SHS).* (Participant 6 - GP - high prevalence area).

HCPs felt that SHS would provide a more streamlined and prompt service than general practice.

> *It’s going to be much more efficient than us…and you’ve got the follow-up already organised.* (Participant 25 - GP - low prevalence area).

#### Capability facilitators

Facilitators for offering an HIV test include having knowledge and training about HIV, an interest in sexual health, experience of working in SHS, and being aware that patients face barriers accessing SHS.

> *I know that for everybody I tell to go to (SHS) some of them won’t go so I think ’let’s just do it’ [test for HIV].* (Participant 16 - GP - high prevalence area)

HCPs suggested that HIV testing should be embedded in existing systems. For example, there was a view by many that HIV testing could and should be part of STI screening or that a well-designed screen prompt (pop-up) on the electronic patient record for ICs could increase HIV testing.

> *If you put in a code that…that could generate a prompt that says something like, have you considered a HIV test in this person, I think that’s – for me, that’s the only way.* (Participant 7 - GP - low prevalence area)

Several participants felt that training for HCPs would facilitate discussions with patients about PrEP.

> *With the right training I would be very happy to.* (Participant 15 - GP - low prevalence area)

#### Opportunity barriers

##### Staff time

Many participants discussed lack of time to carry out a test, often based on assumptions that a lengthy consent process was necessary. There were concerns that in a short consultation there was insufficient time to discuss the test and address patient questions.

> *Time constraints wise is also the other one… you could end up with a whole load of questions which perhaps you don’t have the time to answer in a sort of full way or knowledgeable way.* (Participant 20 - Nurse - low prevalence area).

Participants in high prevalence areas felt that testing according to guidelines would be challenging given the numbers involved. Most participants also felt that there was a lack of time in general practice to initiate, prescribe, or maintain PrEP prescriptions.

#### Opportunity facilitators

Staff suggestions for overcoming opportunity barriers to testing commonly focused on simplifying processes and ensuring that staff are aware that a lengthy consent process is not necessary for HIV tests.

Suggestions for normalising the process, for example, through integrating HIV tests with other blood tests or health checks was mentioned.

> *I wish it was more normalised, for example maybe part of the NHS health check, you know, something very routine, but it almost ought to be added to that really, shouldn’t it?* (Participant 24 - GP - low prevalence area)

Although point of care tests were discussed as an option for increasing testing uptake, the perceived disadvantages of rapid tests (including accuracy and difficulties with finger prick tests) meant that this was rejected as a potential solution.

Conversations about PrEP could be prompted by consultations about sexual health, STIs or by an HIV test.

#### Motivation barriers

#### Reluctance to initiate conversations about HIV

There were several factors involved in reluctance to initiate conversations about HIV. Resources in general practice are stretched and participants described the competing priorities they face. They readily admit that preventative medicine (and HIV prevention) is not a priority.

> *It’s not a priority… just getting through the day of clinical demand for things/symptoms that people have is the priority.* (Participant 14 - GP - high prevalence area)

Some HCPs felt that HIV stigma had an impact on the willingness to discuss testing with patients.

> *I think there’s still a taboo about it…. I don’t have a problem myself asking patients but yeah, I would guess some of my colleagues would certainly find it uncomfortable.* (Participant 15 - GP - low prevalence area)

HIV stigma was recognised as a barrier in small practices where patients might feel uncomfortable interacting with either a member of staff they may have known over many years or one that they may have a personal connection with. Not all staff in smaller practices recognised this as a potential problem though.

> *Most of our practice nurses and GPs have been here for quite a long time…So, the patient was more comfortable speaking to somebody that wasn’t within the practice.* (Participant 5 - Nurse - low prevalence area).

Few HCPs discussed the stigma faced by some patients in their communities if it was known they had had an HIV test. Some HCPs were aware of these barriers to accessing HIV tests and felt that they were hard to address.

> *I have got a patient.. who is [HIV] positive but just says I can’t be… because I think it carries a lot of stigma in her community, it suggests things about your sexuality that…are not true… that’s really hard to combat.* (Participant 10 - GP - high prevalence area)

Motivation to discuss PrEP may also be impacted by consideration of stigma around HIV.

#### Motivation facilitators

Some HCPs were keen to follow testing guidelines if resources or financial incentives could be made available to carry out the additional tests.

> *Without an enhanced service, so money to things and a budget it doesn’t happen.* (Participant 17 - GP - high prevalence area)

HCP and interest holders suggested that normalising the testing process would reduce stigma. For example, if it was a routine activity or part of another routine test so that staff could say to patients ‘this is what we do for everyone’ (Participant 25 - GP - low prevalence area).

A few HCPs mentioned that with appropriate funding they would be interested in initiating PrEP.

> *In terms of PrEP I think it has to be some kind of service where we are funded to provide that.* (Participant 6 - GP - high prevalence area)

However, a lack of system-level commissioning and fragmentation of responsibilities for HIV services were highlighted by interest holder participants who considered that the distribution of funding was a key barrier to increasing general practice activity in HIV prevention.

> *NHS England fund the treatment of HIV or commission it. The local authorities are meant to be doing the prevention… that’s all moving, so the treatment element of HIV is now being delegated to ICBs… but it’s.. been done and still is being done in a regional type basis… we’re not commissioning as a system.* (Participant 27 - Interest Holder - High Prevalence Area)

#### Intervention development and refinement

##### Guiding principles

The intervention’s guiding principles (presented in Supplementary table S5) centre on addressing low HCP motivation to test due to competing priorities and low awareness of HIV prevalence, testing guidelines and PrEP, time pressures to receive and deliver interventions, low self-efficacy to test and discuss PrEP and the perception that SHS may be more appropriate settings for HIV initiatives.

Intervention components with relevant published evidence are below (Supplementary Table S6):

1. Provide short impactful education to address lack of knowledge, skills and confidence around HIV testing and PrEP. To include: training to address lack of clinician knowledge and clinician anxiety associated with discussing HIV testing with patients (50); guidelines, HIV epidemiology and ICs (48); a modified version of a previously tested education intervention (35); education and training to address lack of clinician knowledge and lack of comfort discussing PrEP with patients (59, 60) and to persuade HCPs of the benefits of HIV testing and improving access to PrEP (given barriers to accessing SHS).
2. Introduce systematic ways of identifying when to test according to high and low prevalence areas. To include the provision of the BBV Test Alert (a prompt to test in response to ICs) (18). Electronic health record reminders have been effective in increasing HIV testing (47, 48, 75). Decision aids may improve motivation and capability (76). Using a standardised approach to HIV testing helps to alleviate discomfort (47) and IC testing programmes have been used effectively to increase HIV testing in primary care (50).
3. Simplify the approach to HIV testing. To address time barriers, simplify processes of consent (47), normalise HIV testing using opt-out approaches (without pre-test counselling) which are acceptable to the public (77) and are proven to increase testing (56, 78) and may reduce stigma (47).
4. Communicate HIV policy to patients to raise awareness about the practice’s approach to HIV testing and PrEP and normalise testing processes through regularly communicated information on HIV.
5. Provide leadership and support to implement the intervention through practice champions shown to raise awareness, support and educate colleagues about initiatives to increase HIV testing and to develop ideas and promote shared learning (52).
6. Systematise the signposting of eligible patients to PrEP services.

##### Intervention – co-production and optimisation

Four focus groups comprising 16 HCPs from 8 general practices and 3 focus groups involving 13 members of the public were carried out between November 2023 and April 2024. Table 3 provides demographic information.

**Table 3.**
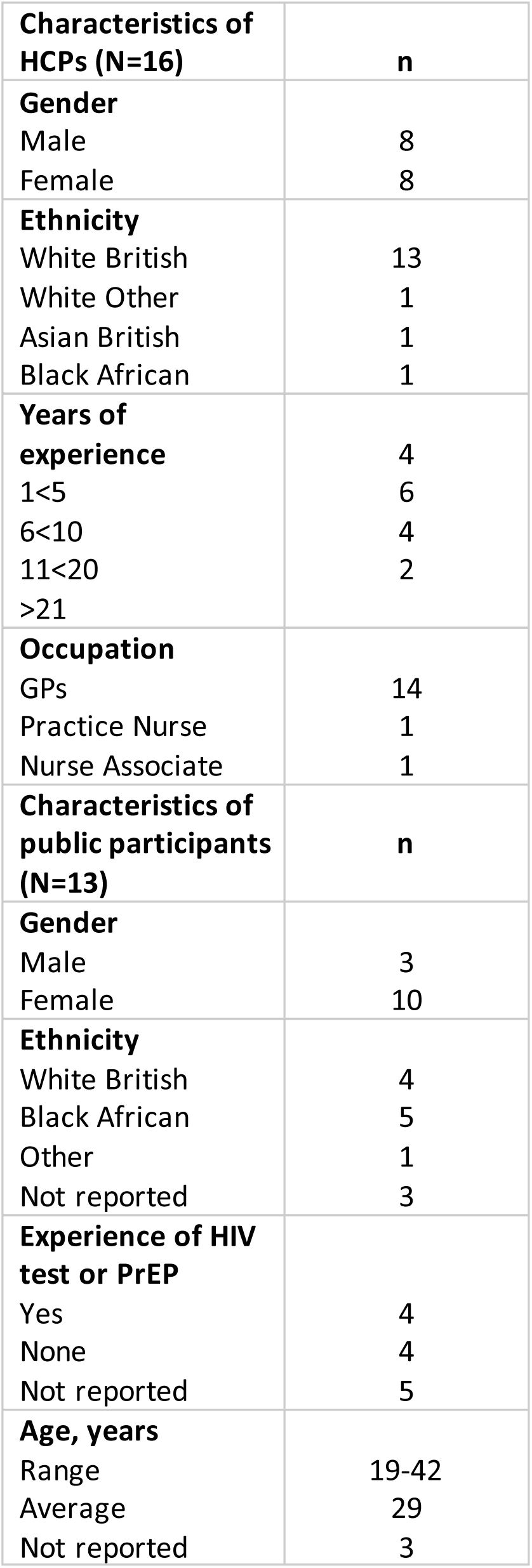
Demographic characteristics of focus groups.

Key focus group findings are presented below and summarised in Box 1.

##### HCP focus group feedback

HCPs viewed many aspects of the intervention positively, for example, simplifying the consent and counselling process. HCPs emphasised the need for training to be brief and in video format to ensure ease of access. The importance of informing all patients about the approach to patient consent using opt-out principles was emphasised. There were some concerns expressed about the potential for patient confusion about an opt-out approach (i.e. not knowing whether an HIV test had been done), particularly non-English speaking patients or those without access to GP digital communication systems. HCPs emphasised the need to ensure that non-English speaking patients received appropriate communication in their own language.

Some expressed concerns about pop-up fatigue regarding the BBV Test Alert (18), however many felt that the advantages of a system which aids decision making would outweigh this. Some felt that these systems would help to normalise HIV and hence reduce stigma.

##### Public focus group feedback

Public feedback was largely positive with some caveats. Participants focused on the importance of communicating the approach to HIV testing effectively and suggested wording that would indicate that testing was universal so that patients would not feel targeted.

Public participants said that it was important that patients were aware that they were being tested and are informed that they can opt-out.

Participants felt that communication information should be simple, engaging and include texting, posters and practice screens. Public participants also emphasised the importance of communicating with non-English speaking people.

Participants wanted access to HIV tests and PrEP to be available at general practices (rather than travelling to sexual health clinics) and this was particularly important for those living in rural communities. These participants felt that GPs should have the knowledge and training to be able to communicate to patients about PrEP.

###### Box 1 HCP and public focus group feedback

|  |
| --- |
| **Focus group findings with participant quotes** |
| **Training should be brief:** *I think, you know, I guess the aim if you could keep it short, succinct, 15/30 minutes then you can give people time for that. But if it starts to get longer than that, you know, that becomes quite a lot of pressure, especially if all of the clinicians need to do it. You know, that would mount up quite a lot.* (HCP Focus group 4 - Participant 1 - GP) |
| **Simplify test processes:** *It's probably much easier to add it on to other tests that we're doing rather than getting every single new patient to come in for a blood test.*<br>(HCP Focus Group 1 - Participant 1 - GP) |
| **Normalise HIV testing:** *If you see someone with thrush a few times then you think, you know, maybe I should do a blood sugar. I think we all routinely think about that now but maybe 30 years ago we didn't and I suspect this will happen with HIV, that it will become a bit more standardised*<br>(HCP Focus group 2 - Participant 2 - GP) |
| **Communicate policy effectively:** *So I think it would be good to have that openness with the patients and always tell them that yes we want to do this test but you have got the uhm... it is the ball is in your court so you can decide whether you want to opt-out or not rather than for them to find out that we done it without their knowledge cos I think once the trust is gone it will be gone.* (HCP Focus Group 3 - Participant 2 - Nurse)<br>*I would like it verbally mentioned and I'd probably be like yeah, yeah, that's fine, but it needs to be clear.* (Public Focus Group 1 - Participant 1)<br>*I also find the television, the screen in the GP very useful for sending information because while we are sitting in the waiting room, that's most of the place that we all look.* (Public Focus Group 3 - Participant 5) |
| **Convenience of opt-out:** *I think it sounds like a really good idea, it would be really convenient for me.* (Public Focus Group 2 - Participant 1) |
| **Convenience of including HIV test as part of STI screening:** *If you've already taken the blood test, you may as well just go for it because at that point we're already in. It's better than having to come back and do another test and keep on doing it. The blood's coming out anyway.* (Public Focus Group 1 - Participant 1) |
| **Electronic health record prompt:** *I think often, with pop-ups though, if they're coming up on every – you know, on loads and loads of patients, you do just tend to switch off.* (HCP Focus Group 1 - Participant 1 - GP)<br>*I think it's probably a good reminder having it that way, and especially having an easy-to-use kind of template as well with clear tick boxes and counselling advice.* (HCP Focus Group 2 - Participant 1 - GP) |
| **Access to PrEP through GP practice would be valued:** *Sexual health clinics is an extra layer of like okay, I've got to find this different special clinic and I've got to find a different set of contact details and work out how the system works and where they are, it's like it's a lot more hassle than the...Like it makes sense for sexual health clinics to offer it but it would make sense for them both to offer it.* (Public Focus Group 2 - Participant 1) |

Fig 2 shows the key design features of the multi-faceted intervention to increase HIV testing and access to PrEP.

**Fig 2.**
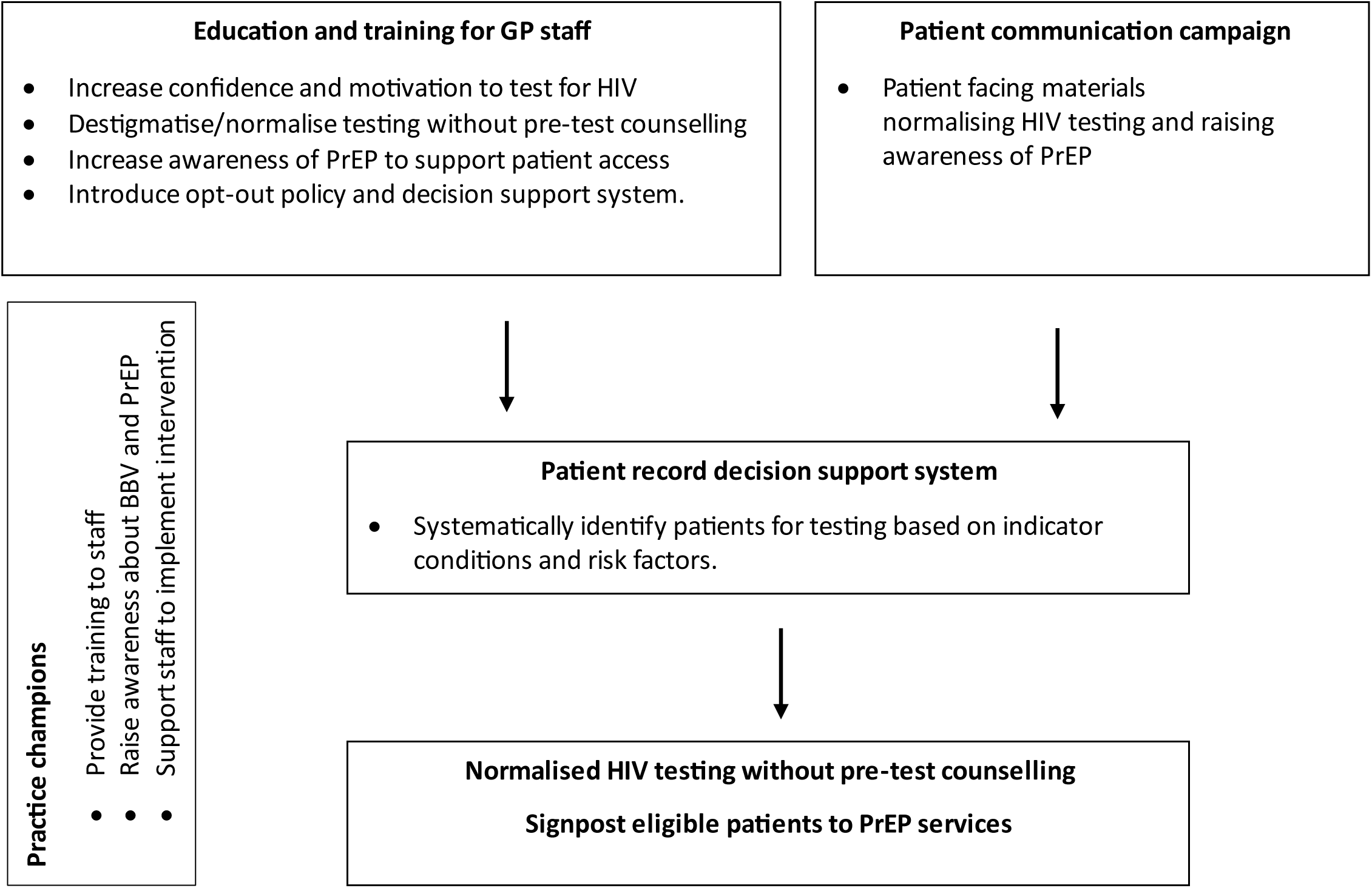
Intervention optimisation – key design features to increase HIV testing and access to PrEP.

## Discussion

### Summary

This paper outlines the development of a multifaceted intervention to support HIV testing and access to PrEP in general practice using the person-based approach. In summary the proposed intervention mapped to elements of the COM-B model involves: 1) HCP education and training to raise awareness of HIV testing and PrEP; 2) a streamlined approach which normalises testing and integrates the process into routine care; 3) a clinical decision support application embedded within the electronic health record; 4) a patient communication campaign; and 5) the introduction of practice champions to support the intervention implementation.

Research is now needed to explore the feasibility and the effectiveness of the intervention. If efficacy can be demonstrated, this intervention has the potential to support the goal to eliminate new cases of HIV by 2030.

### Strengths and limitations

A strength of this research is the use of a co-design approach to develop materials, integrating insights from academic literature, qualitative interviews, behavioural science theory and patient and public involvement. This allowed us to gain a comprehensive understanding of the context and differing perspectives identify a wide range of barriers to testing and supporting access to PrEP and collaboratively discuss potentially accessible, acceptable, persuasive and feasible approaches to address them. We aimed to be as inclusive as possible and used various approaches to collect data from a range of practices, locations, HCPs and members of the public. Such widescale involvement may help optimise the acceptability, accessibility and feasibility of the intervention.

Despite best efforts, key voices may have been missed. Although we were able to hear from people who do not speak English as a first language, this group were very health literate, and we were unable to hear from those who do not read or write in English. Given the need for communication with the public about opt-out testing, it is critical that we engage with the latter before the intervention is implemented.

### Comparison with existing literature

In line with previous research (47, 79) we found testing behaviour to be influenced by HCP’s capability, motivation and opportunity to offer tests. Indeed, a key theme from the planning stage highlights that most HCPs are not aware of the local HIV prevalence, and did not know what this meant in terms of the HIV testing guidelines (37, 47–49).

To increase testing in accordance with guidelines, it was clear that there is a need to raise awareness of the testing guidelines and HIV prevalence in local area. The findings highlighted how educational materials should be delivered to work within general practice (e.g., must be short, online and be delivered flexibly). Intervention content targeting knowledge also needed to go beyond the provision of information and must also include prompts or reminders when patients present with ICs (56).

A few HCPs interviewed in the study identified pop-ups as a way to facilitate testing according to ICs and in focus groups HCPs found this an acceptable to increase the identification of people living with HIV and reduce late diagnosis. Currently, testing by ICs are unevenly and insufficiently implemented across conditions and healthcare settings (56, 80). However, interventions using ICs to guide testing have increased testing rates and diagnoses (50, 55). For example, implementation of a normative illness script-based decision-making model, called BBV Test Alert, prompts HCPs to add a BBV test in response to ICs. A pilot of the Test Alert led to five times more HIV testing and three times more hepatitis B and hepatitis C testing. While the pilot demonstrated the Test Alert’s feasibility, research is needed to determine its effectiveness and cost-effectiveness (45). Testing returned to baseline levels potentially due to alert fatigue or because they had reached the population requiring testing. However, the model also increased consultation number and time (18), thus this approach in isolation may not be appealing to HCPs.

In addition to barriers relating to knowledge of HIV and PrEP, motivation and capacity for testing was also low. For educational campaigns to be effective they needed to be paired with approaches for simplifying the testing process. Indeed, many HCPs were reluctant to offer a test or initiate conversations about PrEP because of a lack of time. This was in part driven by the perception that it would be necessary to engage in lengthy conversation and pre-test counselling with the patient before a test could be offered. Many HCPs assumed that patients would have concerns and questions if they were to discuss HIV. Patients who may benefit from PrEP were also assumed by HCPs to know where they could access it, whereas data on PrEP access suggests there is a need to improve awareness of PrEP among key groups and (81) barriers to access through SHS exist (6).

Motivation was reduced when HIV was perceived as a low priority, as even in high prevalence areas, 2 per 1000 practice population are small numbers. Whilst numerous approaches to increasing motivation and reducing stigma were identified, the most acceptable approach needed to reduce rather than extend consultation time. Opt-out approaches to patient consent to testing (82) - in which a person is informed that HIV testing is routine/standard of care and they actively decline if they do not wish to be tested for HIV - may help address these barriers to testing and may help reduce the time to discuss testing, reduce stigma (47) and normalise testing (83). Opt-out approaches have been successfully adopted within antenatal services and showed promise for new patient registrations in an area with high HIV prevalence (16). BBV opt-out testing in emergency departments within very high prevalence areas has led to increased testing in England (84). Using a combination of previous successful strategies including IC guided testing, training and education and opt-out approaches could be a feasible and acceptable approach to increase HIV testing (56).

Structural barriers noted by interest holder participants (for example, the fragmentation of funding responsibilities for HIV services) has made it challenging to deliver coordinated and planned care across the system to meet people’s needs (85). This has led to disparities in types of HIV tests and testing opportunities across the system (86). New BASHH/BHIVA guidance (under consultation) recommends expanding PrEP provision to beyond SHS including primary care (with regulation from SHS) (87) and there are plans to explore piloting HIV PrEP referrals from primary care to specialist SHS (81). General practice interventions to support access to PrEP are therefore timely.

### Implications for research and/or practice

Whilst this research has led to the development of an intervention that is potentially acceptable to HCPs and patients, research is needed to test the real -world feasibility and effectiveness of the intervention for increasing HIV testing and supporting access to PrEP. Although there is promising evidence for each aspect of our co-produced intervention in isolation, there is a need for evaluation of the intervention as a whole. However, the wider context of structural fragmentation of responsibility for HIV will also need to be addressed to facilitate increased HIV testing and access to PrEP.

## Supporting information

Supplemental Box, Tables and Figure

## Data Availability

Data will be available at the University of Bristol data repository data.bris. Data access will be restricted to bona fide researchers for ethically approved research and subject to approval by the University's Data Access Committee.

## Acknowledgments

We thank Lydia Holt for assisting with the scoping review. We are grateful to the PPI panel for their valuable input and to those who participated in the study.

Jo Kesten and Sarah Denford are joint senior authors.

## Ethical approval

This study received ethical approval from the University of Bristol’s Faculty of Health Sciences Research Ethics Committee (Ref: 12240), and Health Research Authority approval to work with local NHS services (IRAS 321564, Date 16/01/2023).

## Data

Data will be available at the University of Bristol data repository, data.bris. Data access will be restricted to bona fide researchers for ethically approved research and subject to approval by the University’s Data Access Committee.

## Supporting information

**Supplementary Box S1 – Example Medline search strategies**

**Supplementary Box S2 – Interview topic guide**

**Supplementary Table S3 – Behavioural analysis planning table**

**Supplementary Figure S4 – Logic Model**

**Supplementary Table S5 – Guiding Principles**

**Supplementary Table S6 – Developing the intervention**

## References

1. UNAIDS. Fast-Track - Ending the AIDS epidemic by 2030 2014 [Available from: https://www.unaids.org/en/resources/documents/2014/JC2686_WAD2014report.

2. Department of Health and Social Care. Towards Zero - An action plan towards ending HIV transmission, AIDS and HIV-related deaths in England - 2022 to 2025. 2021.

3. National Institute for Health and Care Excellence (NICE). HIV testing: encouraging uptake. Quality standard [QS157] 2017 [

4. National Institute for Health and Care Excellence (NICE). Reducing sexually transmitted infections. NICE guideline [NG221]. Recommendations for research. 2022 [Available from: https://www.nice.org.uk/guidance/ng221/chapter/Recommendations-for-research.

5. UKHSA. Research and analysis. HIV Action Plan monitoring and evaluation framework 2024 report 2024 [Available from: https://www.gov.uk/government/publications/hiv-monitoring-and-evaluation-framework/hiv-action-plan-monitoring-and-evaluation-framework-2024-report.

6. UKHSA. Official Statistics. HIV testing, PrEP, new HIV diagnoses and care outcomes for people accessing HIV services: 2024 report 2024 [Available from: https://www.gov.uk/government/statistics/hiv-annual-data-tables/hiv-testing-prep-new-hiv-diagnoses-and-care-outcomes-for-people-accessing-hiv-services-2024-report.

7. HIV Prevention England. GPs and primary care professionals 2018 [Available from: https://www.hivpreventionengland.org.uk/evidence-and-guidance/gps-and-primary-care-professionals/.

8. Girardi E, Sabin CA, Monforte AdA. Late diagnosis of HIV infection: Epidemiological features, consequences and strategies to encourage earlier testing. Jaids-Journal of Acquired Immune Deficiency Syndromes. 2007;46:S3–S8.

9. May M, Gompels M, Delpech V, Porter K, Post F, Johnson M, et al. Impact of late diagnosis and treatment on life expectancy in people with HIV-1: UK Collaborative HIV Cohort (UK CHIC) Study. British Medical Journal. 2011;343(d6016).

10. Beck EJ, Mandalia S, Sangha R, Sharott P, Youle M, Baily G, et al. The Cost-Effectiveness of Early Access to HIV Services and Starting cART in the UK 1996-2008. PLoS One. 2011;6(12).

11. Trickey A, Sabin CA, Burkholder G, Crane H, d’Arminio Monforte A, Egger M, et al. Life expectancy after 2015 of adults with HIV on long-term antiretroviral therapy in Europe and North America: a collaborative analysis of cohort studies. Lancet HIV. 2023;10(5):e295–e307.

12. Rogers A, Bruun T, Cambiano V, Vernazza P, Estrada V, Van Lunzen J, et al. HIV Transmission Risk Through Condomless Sex If HIV+ Partner On Suppressive ART: PARTNER study (CROI abstract 153LB). Abstracts from the 2014 Conference on Retroviruses and Opportunistic Infections. Top Antivir Med. 2014;22((e-1)):34.

13. Baggaley RF, Irvine MA, Leber W, Cambiano V, Figueroa J, McMullen H, et al. Cost-effectiveness of screening for HIV in primary care: a health economics modelling analysis. Lancet HIV. 2017;4(10):e465–e74.

14. Kall MM, Smith RD, Delpech VC. Late HIV diagnosis in Europe: a call for increased testing and awareness among general practitioners. Eur J Gen Pract. 2012;18(3):181–6.

15. Rayment M, Thornton A, Mandalia S, Elam G, Atkins M, Jones R, et al. HIV Testing in Non-Traditional Settings – The HINTS Study: A Multi-Centre Observational Study of Feasibility and Acceptability. PLOS ONE. 2012;7(6):e39530.

16. Health Protection Agency. Time to test for HIV: expanding HIV testing in healthcare and community services in England. Final report, 2011. 2011.

17. Elmahdi R, Gerver SM, Gomez Guillen G, Fidler S, Cooke G, Ward H. Low levels of HIV test coverage in clinical settings in the U.K.: a systematic review of adherence to 2008 guidelines. Sexually Transmitted Infections. 2014;90(2):119–24.

18. Chadwick D, Forbes G, Lawrence C, Lorrimer S, van Schaik P. Using an electronic health record alert to prompt blood-borne virus testing in primary care. AIDS. 2021;35(11):1845–50.

19. Davies CF, Kesten JM, Gompels M, Horwood J, Crofts M, Billing A, et al. Evaluation of an educational intervention to increase HIV-testing in high HIV prevalence general practices: a pilot feasibility stepped-wedged randomised controlled trial. BMC Family Practice. 2018;19(1):195.

20. Burns FM, Johnson AM, Nazroo J, Ainsworth J, Anderson J, Fakoya A, et al. Missed opportunities for earlier HIV diagnosis within primary and secondary healthcare settings in the UK. AIDS. 2008;22(1):115–22.

21. Read P, Armstrong-James D, Tong CYW, Fox J. Missed opportunities for HIV testing-a costly oversight. Qjm-an International Journal of Medicine. 2011;104(5):421–4.

22. Sudarshi D, Pao D, Murphy G, Parry J, Dean G, Fisher M. Missed opportunities for diagnosing primary HIV infection. Sexually transmitted infections. 2008;84(1):14–6.

23. Watipa UKHSA. HIV Lens 2025 [Available from: https://www.hiv-lens.org/.

24. National Institute of Health and Care Excellence (NICE). HIV testing: increasing uptake among people who may have undiagnosed HIV. NIC guideline [NG60] 2016 [Available from: https://www.nice.org.uk/guidance/ng60.

25. British HIV Association (BHIVA). British HIV Association/British Association for Sexual Health and HIV/British Infection Association Adult HIV Testing Guidelines 2020. 2020.

26. Smith C. HIV: low prevalence is no excuse for not testing. British Journal of General Practice. 2011;61(588):436–7.

27. Gompels M, Michael S, Davies C, Jones T, Macleod J, May M. Trends in HIV testing in the UK primary care setting: a 15-year retrospective cohort study from 2000 to 2015. BMJ Open. 2019;9(11):e027744.

28. Stankevitz K, Nhamo D, Murungu J, Ridgeway K, Mamvuto T, Lenzi R, et al. Test and Prevent: Evaluation of a Pilot Program Linking Clients With Negative HIV Test Results to Pre-exposure Prophylaxis in Zimbabwe. Global Health: Science and Practice. 2021;9(1):40–54.

29. Rodger AJ, Cambiano V, Bruun T, Vernazza P, Collins S, Degen O, et al. Risk of HIV transmission through condomless sex in serodifferent gay couples with the HIV-positive partner taking suppressive antiretroviral therapy (PARTNER): final results of a multicentre, prospective, observational study. The Lancet. 2019;393(10189):2428–38.

30. Brunner P, Brunner K, Kübler D. The Cost-Effectiveness of HIV/STI Prevention in High-Income Countries with Concentrated Epidemic Settings: A Scoping Review. AIDS and Behavior. 2022.

31. Grimshaw C, Estcourt CS, Nandwani R, Yeung A, Henderson D, Saunders JM. Implementation of a national HIV pre-exposure prophylaxis service is associated with changes in characteristics of people with newly diagnosed HIV: a retrospective cohort study. Sex Transm Infect. 2022;98(1):53–7.

32. Sullivan AK, Saunders J, Desai M, Cartier A, Mitchell HD, Jaffer S, et al. HIV pre-exposure prophylaxis and its implementation in the PrEP Impact Trial in England: a pragmatic health technology assessment. The Lancet HIV. 2023;10(12):e790–e806.

33. HIV Prevention England. HIV in Primary Care: An introduction to PrEP 2018 [Available from: https://www.hivpreventionengland.org.uk/wp-content/uploads/2019/02/Introduction-to-PrEP.pdf.

34. Pinto RM, Berringer KR, Melendez R, Mmeje O. Improving PrEP Implementation Through Multilevel Interventions: A Synthesis of the Literature. AIDS and Behavior. 2018;22(11):3681–91.

35. Kesten JM, Davies CF, Gompels M, Crofts M, Billing A, May MT, et al. Qualitative evaluation of a pilot educational intervention to increase primary care HIV-testing. BMC Family Practice. 2019;20(1):74.

36. Bailey AC, Dean G, Hankins M, Fisher M. Attending an STI Foundation course increases chlamydia testing in primary care, but not HIV testing. Int J STD AIDS. 2008;19(9):633–4.

37. Mahendran P, Soni S, Goubet S, Saunsbury E, Roberts J, Fisher M. Testing initiatives increase rates of HIV diagnosis in primary care and community settings: an observational single-centre cohort study. PLoS One. 2015;10(4):e0124394.

38. Pillay TD, Mullineux J, Smith CJ, Matthews P. Unlocking the potential: longitudinal audit finds multifaceted education for general practice increases HIV testing and diagnosis. Sex Transm Infect. 2013;89(3):191–6.

39. Bogers S, Schim van der Loeff M, Boyd A, van Dijk N, Geerlings S, van Bergen J, et al. Improving provider-initiated testing for HIV and other STI in the primary care setting in Amsterdam, the Netherlands: Results from a multifaceted, educational intervention programme. Plos one. 2023;18(3):e0282607.

40. Shojania KG, Jennings A, Ramsay CR, Grimshaw JM, Kwan JL, Lo L. The effects of on-screen, point of care computer reminders on processes and outcomes of care. Cochrane Database of Systematic Reviews. 2009(3).

41. Forsetlund L, O’Brien MA, Forsén L, Mwai L, Reinar LM, Okwen MP, et al. Continuing education meetings and workshops: effects on professional practice and healthcare outcomes. Cochrane Database of Systematic Reviews. 2021(9).

42. Yardley L, Morrison L, Bradbury K, Muller I. The Person-Based Approach to Intervention Development: Application to Digital Health-Related Behavior Change Interventions. J Med Internet Res. 2015;17(1):e30.

43. Horwood J, Pithara C, Lorenc A, Kesten JM, Murphy M, Turner A, et al. The experience of conducting collaborative and intensive pragmatic qualitative (CLIP-Q) research to support rapid public health and healthcare innovation. Frontiers in Sociology. 2022;7.

44. Michie S, van Stralen MM, West R. The behaviour change wheel: A new method for characterising and designing behaviour change interventions. Implementation Science. 2011;6(1):42.

45. Peters MDJ GC, McInerney P, Munn Z, Tricco AC, Khalil, H. Scoping Reviews (2020). 2024. In: JBI Manual for Evidence Synthesis [Internet]. Available from: https://synthesismanual.jbi.global/.

46. Department of Health and Social Care. Sexual and Reproductive Health Profiles 2024 [Available from: https://fingertips.phe.org.uk/profile/sexualhealth.

47. Deblonde J, Van Beckhoven D, Loos J, Boffin N, Sasse A, Nöstlinger C, et al. HIV testing within general practices in Europe: a mixed-methods systematic review. BMC Public Health. 2018;18(1):1191.

48. Tan K, Black BP. A Systematic Review of Health Care Provider-Perceived Barriers and Facilitators to Routine HIV Testing in Primary Care Settings in the Southeastern United States. Journal of the Association of Nurses in AIDS Care. 2018;29(3):357–70.

49. Serag H, Clark I, Naig C, Lakey D, Tiruneh YM. Financing Benefits and Barriers to Routine HIV Screening in Clinical Settings in the United States: A Scoping Review. International Journal of Environmental Research & Public Health [Electronic Resource]. 2022;20(1):27.

50. Desai S, Tavoschi L, Sullivan AK, Combs L, Raben D, Delpech V, et al. HIV testing strategies employed in health care settings in the European Union/European Economic Area (EU/EEA): evidence from a systematic review. HIV Medicine. 2020;21(3):163–79.

51. Loos J, Manirankunda L, Hendrickx K, Remmen R, Nöstlinger C. HIV testing in primary care: feasibility and acceptability of provider initiated HIV testing and counseling for sub-Saharan African migrants. AIDS Education and Prevention. 2014;26(1):81–93.

52. Fraser AC, C. Karamanos, A. Service Evaluation of the Elton John AIDS Foundation’s Zero HIV Social Impact Bond intervention in South London: An investigation into the implementation and sustainability of activities and system changes designed to bring us closer to an AIDS free future. Final Report, King’s College London. 2022.

53. Davis M, Hoskins K, Phan M, Hoffacker C, Reilly M, Fugo PB, et al. Screening Adolescents for Sensitive Health Topics in Primary Care: A Scoping Review. Journal of Adolescent Health. 2022;70(5):706–13.

54. Chavez PRG, Wesolowski LG, Peters PJ, Johnson CH, Nasrullah M, Oraka E, et al. How well are US primary care providers assessing whether their male patients have male sex partners? Preventive Medicine. 2018;107:75–80.

55. Davies C, May M, Gompels M. Use and effectiveness of HIV indicator conditions in guiding HIV testing: a review of the evidence. International STD Research and Reviews. 2017;6(2):36373.

56. Bogers S, Hulstein S, van der Loeff MS, de Bree G, Reiss P, van Bergen J, et al. Current evidence on the adoption of indicator condition guided testing for HIV in western countries: A systematic review and meta-analysis. EClinicalMedicine. 2021;35.

57. Hillis A, Germain J, Hope V, McVeigh J, Van Hout MC. Pre-exposure prophylaxis (PrEP) for HIV prevention among men who have sex with men (MSM): a scoping review on PrEP service delivery and programming. AIDS and Behavior. 2020;24:3056–70.

58. Conley C, Johnson R, Bond K, Brem S, Salas J, Randolph S. US Black cisgender women and pre-exposure prophylaxis for human immunodeficiency virus prevention: A scoping review. Women’s Health. 2022;18:17455057221103098.

59. Hoornenborg E, Krakower DS, Prins M, Mayer KH. Pre-exposure prophylaxis for MSM and transgender persons in early adopting countries. Aids. 2017;31(16):2179–91.

60. Pleuhs B, Quinn KG, Walsh JL, Petroll AE, John SA. Health Care Provider Barriers to HIV Pre-Exposure Prophylaxis in the United States: A Systematic Review. AIDS Patient Care & Stds. 2020;34(3):111–23.

61. Zhang C, McMahon J, Fiscella K, Przybyla S, Braksmajer A, LeBlanc N, et al. HIV pre-exposure prophylaxis implementation cascade among health care professionals in the United States: implications from a systematic review and meta-analysis. AIDS patient care and STDs. 2019;33(12):507–27.

62. Matacotta JJ, Rosales-Perez FJ, Carrillo CM. HIV Preexposure Prophylaxis and Treatment as Prevention - Beliefs and Access Barriers in Men Who Have Sex With Men (MSM) and Transgender Women: A Systematic Review. Journal of Patientcentered Research & Reviews. 2020;7(3):265–74.

63. Mayer KH, Agwu A, Malebranche D. Barriers to the wider use of pre-exposure prophylaxis in the United States: a narrative review. Advances in therapy. 2020;37:1778–811.

64. Silapaswan A, Krakower D, Mayer KH. Pre-Exposure Prophylaxis: A Narrative Review of Provider Behavior and Interventions to Increase PrEP Implementation in Primary Care. Journal of General Internal Medicine. 2017;32:192–8.

65. Wang Y, Mitchell JW, Zhang C, Liu Y. Evidence and implication of interventions across various socioecological levels to address pre-exposure prophylaxis uptake and adherence among men who have sex with men in the United States: a systematic review. AIDS Research & Therapy [Electronic Resource]. 2022;19(1):28.

66. Zablotska IB, O’Connor CC. Preexposure prophylaxis of HIV infection: the role of clinical practices in ending the HIV epidemic. Current HIV/AIDS Reports. 2017;14:201–10.

67. Bradley E, Forsberg K, Betts JE, DeLuca JB, Kamitani E, Porter SE, et al. Factors Affecting Pre-Exposure Prophylaxis Implementation for Women in the United States: A Systematic Review. Journal of Women’s Health. 2019;28(9):1272–85.

68. Li DH, Benbow N, Keiser B, Mongrella M, Ortiz K, Villamar J, et al. Determinants of implementation for HIV pre-exposure prophylaxis based on an updated consolidated framework for implementation research: a systematic review. JAIDS Journal of Acquired Immune Deficiency Syndromes. 2022;90(S1):S235–S46.

69. Siegler AJ, Steehler K, Sales JM, Krakower DS. A review of HIV pre-exposure prophylaxis streamlining strategies. Current HIV/AIDS Reports. 2020;17:643–53.

70. Cooper RL, Juarez PD, Morris MC, Ramesh A, Edgerton R, Brown LL, et al. Recommendations for Increasing Physician Provision of Pre-Exposure Prophylaxis: Implications for Medical Student Training. Inquiry. 2021;58:469580211017666.

71. Allen E, Gordon A, Krakower D, Hsu K. HIV preexposure prophylaxis for adolescents and young adults. Current Opinion in Pediatrics. 2017;29(4):399–406.

72. Saberi P, Mehtani NJ, Sayegh A, Camp CE, Chu C. Understanding HIV Pre-Exposure Prophylaxis Questions of U.S. Health Care Providers: Unique Perspectives from the PrEPline Clinical Teleconsultation Service. Telemedicine Journal & E Health. 2022;8:08.

73. Yusuf H, Fields E, Arrington-Sanders R, Griffith D, Agwu AL. HIV Preexposure Prophylaxis among Adolescents in the US: A Review. JAMA Pediatrics. 2020;174:1102–8.

74. Turner L, Roepke A, Wardell E, Teitelman AM. Do You PrEP? A Review of Primary Care Provider Knowledge of PrEP and Attitudes on Prescribing PrEP. Journal of the Association of Nurses in AIDS Care. 2018;29(1):83–92.

75. Romero RA, Klausner JD, Marsch LA, Young SD. Technology-delivered intervention strategies to bolster HIV testing. Current HIV/AIDS Reports. 2021;18(4):391–405.

76. Le Bonniec A, Sun S, Andrin A, Dima AL, Letrilliart L. Barriers and Facilitators to Participation in Health Screening: an Umbrella Review Across Conditions. Prevention science : the official journal of the Society for Prevention Research. 2022;23:1115–42.

77. Glew S, Pollard A, Hughes L, Llewellyn C. Public attitudes towards opt-out testing for HIV in primary care: a qualitative study. British Journal of General Practice. 2014;64(619):e60–e6.

78. Soh QR, Oh LY, Chow EP, Johnson CC, Jamil MS, Ong JJ. HIV testing uptake according to opt-in, opt-out or risk-based testing approaches: a systematic review and meta-analysis. Current HIV/AIDS Reports. 2022;19(5):375–83.

79. Bagchi AD, Davis T. Clinician Barriers and Facilitators to Routine HIV Testing: A SystematicReview of the Literature. J Int Assoc Provid AIDS Care. 2020;19:2325958220936014-.

80. Bogers SJ, Schim van der Loeff MF, van Dijk N, Groen K, Groot Bruinderink ML, de Bree GJ, et al. Rationale, design and initial results of an educational intervention to improve provider-initiated HIV testing in primary care. Fam Pract. 2021;38(4):441–7.

81. Department of Health and Social Care. Independent report: Roadmap for meeting the PrEP needs of those at significant risk of HIV 2024 [Available from: https://www.gov.uk/government/publications/roadmap-for-meeting-the-prep-needs-of-those-at-significant-risk-of-hiv/roadmap-for-meeting-the-prep-needs-of-those-at-significant-risk-of-hiv#annex-a-recommendations-of-the-prep-access-and-equity-task-and-finish.

82. Chadwick D, Palfreeman, A, Sullivan, A, Peto, T, Cromarty, B, Hill-Tout, R, Hunter, L, McQuillan, O, Phillips, M, Rodger, A, Smith, S, van Halsema, C, Young, E. British HIV Association (BHIVA), British Association for Sexual Health and HIV (BASHH), British Infection Association (BIA) and Royal College of Emergency Medicine (RCEM) joint working group: rapid guidance on opt-out blood-borne virus testing in high-prevalence and extremely high prevalence acute medical settings and emergency departments.

83. British HIV Association (BHIVA). British HIV Association (BHIVA), British Association for Sexual Health and HIV (BASHH), British Infection Association (BIA) and Royal College of Emergency Medicine (RCEM) joint working group: rapid guidance on opt-out blood-borne virus testing in high-prevalence and extremely highprevalence acute medical settings and emergency departments. 2024.

84. Adebayo-Clement O, Allison, S, Brown, A, Buitendam, E, Clancy, I, Desai, M, Edney, J, Hill-Tout, R, Hindle, S, Hopkins, S, Horwood, J, Humphreys, C, Lester, J, Lowndes, K, Mandal, S, Martin, V, May, T, Moore, H, Mou, D, Murdoch, S, Newbigging-Lister, A, Roche, R, Simmons, Tabor, R, Threadgold, G. Emergency department bloodborne virus opt-out testing: 12-month interim report 2023 2023 [Available from: https://www.gov.uk/government/publications/bloodborne-viruses-opt-out-testing-in-emergency-departments/emergency-department-bloodborne-virus-opt-out-testing-12-month-interim-report-2023.

85. Baylis AB, D. Anderson, J. Jabbal, J. Ross, S. The future of HIV services in England: shaping the response to changing needs. The King’s Fund; 2017.

86. Coukan F, Murray KK, Papageorgiou V, Lound A, Saunders J, Atchison C, et al. Barriers and facilitators to HIV pre-exposure prophylaxis (prep) in specialist sexual health services in the United kingdom: a systematic review using the prep care continuum. HIV medicine. 2023;24(8):893–913.

87. BASHH/BHIVA. BASHH/BHIVA guidelines on the use of HIV pre-exposure prophylaxis (PrEP)(2024) 2024 [Available from: https://www.bashh.org/_userfiles/pages/files/draft_bashh_bhiva_prep_guidelines_240924_v30_final.pdf.

