## Supplemental Box, Tables and Figure for "Developing the PATH-GP (Prevention and Testing for HIV in General Practice) intervention: a Person-Based Approach intervention development study to increase HIV testing and PrEP access"

**Supplementary Box S1 – Example Medline search strategies**

**HIV testing in primary care**

1 exp HIV/ or exp HIV Infections/347319
2 Mass Screening/ 115584
3 Point-of-Care Testing/ or Point-of-Care Systems/ 19366
4 2 or 3 134527
5 1 and 4 8261
6 ((HIV or HIV-1* or HIV1* or HIV-2* or HIV2* or human immunodeficien* vir* or human immunedeficien* vir* or human immun* deficien* vir* or acquired immunodeficien* syndrom* or acquired immunedeficien* syndrome* or immun* deficien* syndrom*) adj3 (test* or screen*)).tw,kf. 33231
7 5 or 6 36260
8 physician-patient relations/ 76018
9 primary health care/ or *"continuity of patient care"/ or *patient handoff/ 101251
10 physicians, family/ 17146
11 general practice/ or family practice/ 78172
12 *family health/7810
13 (primary adj (care or health care or healthcare)).tw,kf. 147719
14 (GP or "GP’s" or generalist*).tw,kf. 80700
15 ((general or family or nurs*) adj (practice* or practitioner*)).tw,kf. 135287
16 nurse practitioners/ 18775
17 primary care nursing/ 565
18 family nursing/ 1564
19 *community health centers/ or *substance abuse treatment centers/ or *community mental health centers/9614
20 ((family or community or practi*) adj (medic* or doctor* or physician* or health* or nurs*)).tw,kf. 102944
21 or/8-20 537646
22 7 and 21 2188
23 "systematic review"/ or meta-analysis/ or network meta-analysis/ 303433
24 (systematic or structured or evidence or trials or studies).ti. and ((review or overview or look or examination or update* or summary).ti. or review.pt.) 314052
25 (0266-4623 or 1469-493X or 1366-5278 or 1530-440X or 2046-4053).is. 20829
26 meta-analysis.pt. or (meta-analys* or meta analys* or metaanalys* or meta synth* or meta-synth* or metasynth*).ti,ab,kf,hw. 294138
27 ((systematic or meta) adj2 (analys* or review)).ti,kf. or ((systematic* or quantitativ* or methodologic*) adj5 (review* or overview*)).ti,ab,kf,sh. or ((quantitativ* or qualitativ*) adj5 synth*).ti,ab,kf,hw. 410237
28 (integrative research review* or research integration).tw. or scoping review?.ti,kf. or (review.ti,kf,pt. and (trials as topic or studies as topic).hw.) or (evidence adj3 review*).ti,ab,kf. 236243
29 review.pt. and (medline or medlars or embase or pubmed or scisearch or psychinfo or psycinfo or psychlit or psyclit or cinahl or electronic database* or bibliographic database* or computeri#ed database* or online database* or pooling or pooled or mantel haenszel or peto or dersimonian or der simonian or fixed effect or (hand adj2 search*) or (manual* adj2 search*)).tw,hw. 207027
30 ((systematic* or quantitativ* or qualitativ* or methodologic* or evidence) adj5 (review* or overview*)).tw. 387113
31 (umbrella review or "review of reviews").tw. 1979
32 (rapid review or mixed method? review or scoping review or literature review).tw. 141432

33 or/23-32 867960
34 22 and 33 76
35 ((HIV or HIV-1* or HIV1* or HIV-2* or HIV2* or human immunodeficien* vir* or human immunedeficien* vir* or human immun* deficien* vir* or acquired immunodeficien* syndrom* or acquired immunedeficien* syndrome* or immun* deficien* syndrom*) adj3 (test* or screen*)).ti. 11322
36 21 and 35 846
37 limit 36 to "review articles" 29
38 34 or 37 93
39 afghanistan/ or exp africa/ or albania/ or andorra/ or antarctic regions/ or argentina/ or exp asia, central/ or exp asia, northern/ or exp asia, southeastern/ or exp atlantic islands/ or bahrain/ or bangladesh/ or Bhutan/ or bolivia/ or borneo/ or "bosnia and herzegovina"/ or brazil/ or bulgaria/ or exp central america/ or exp china/ or "commonwealth of independent states"/ or croatia/ or "democratic people's republic of korea"/ or ecuador/ or gibraltar/ or guyana/ or exp india/ or indonesia/ or iran/ or iraq/ or jordan/ or kosovo/ or kuwait/ or lebanon/ or liechtenstein/ or macau/ or "macedonia (republic)"/ or exp melanesia/ or moldova/ or monaco/ or mongolia/ or montenegro/ or nepal/ or netherlands antilles/ or new guinea/ or oman/ or pakistan/ or paraguay/ or peru/ or philippines/ or qatar/ or "republic of belarus"/ or romania/ or exp russia/ or saudi arabia/ or serbia/ or sri lanka/ or suriname/ or syria/ or taiwan/ or exp transcaucasia/ or ukraine/ or uruguay/ or united arab emirates/ or exp ussr/ or venezuela/ or yemen/ or sub-saharan africa*.ti,kf. 1314584
40 "organisation for economic co-operation and development"/507
41 australasia/ or exp australia/ or austria/ or exp baltic states/ or belgium/ or exp canada/ or chile/ or czech republic/ or colombia/ or europe/ or exp france/ or exp germany/ or greece/ or hungary/ or ireland/ or israel/ or exp italy/ or exp japan/ or korea/ or luxembourg/ or mexico/ or netherlands/ or new zealand/ or north america/ or poland/ or portugal/ or exp "republic of korea"/ or exp "scandinavian and nordic countries"/ or slovakia/ or slovenia/ or spain/ or switzerland/ or turkey/ or exp united kingdom/ or exp united states/3464164
42 european union/ 17538
43 developed countries/ 21307
44 40 or 41 or 42 or 43 3479953
45 39 not 44 1222949
46 38 not 45 75
47 (advisory committees/ or consensus/ or guideline/ or practice guideline/ or health policy/ or international health regulations/ or (advisory committee or advisory group or guideline or consensus statement).ti,ab,kf. or (expert adj (review or recommendation)).mp. or guideline.pt.) and (NHS or NICE or "National Institute for Clinical Excellence" or "National Institute for Health and Care Excellence" or UK or GB or Great Britain or England or Ireland or Scotland or Wales).mp. 10662
48 35 and 47 29
49 limit 48 to "review articles" 4
50 46 or 49 79

**PrEP in primary care**

1 exp HIV/ 106287
2 exp HIV Infections/ 310528
3 (HIV or HIV-1* or HIV1* or HIV-2* or HIV2* or human immunodeficien* vir* or human immunedeficien* vir* or human immun* deficien* vir* or acquired immunodeficien* syndrom* or acquired immunedeficien* syndrome* or immun* deficien* syndrom*).mp. 459042
4 AIDS related.mp. 31557

5 or/1-4 464692
6 Pre-Exposure Prophylaxis/ 4367
7 (PrEP or pre expos* prophyla* or preexpos* prophyla*).mp. 9976
8 *Anti-Retroviral Agents/tu and (prevent* or prophyla*).af. 1618
9 ((antiretrovir* or anti-retrovir* or HIV) adj prophyla*).mp. 723
10 impact trial.mp. 180
11 or/6-10 12244
12 5 and 11 8411
13 primary health care/ or "continuity of patient care"/ or patient handoff/ or transition to adult care/ 111139
14 Physicians, Family/ 17078
15 general practice/ or family practice/ 77973
16 *Family Health/ 7804
17 (primary adj2 (care or health*)).tw,kf. 178595
18 (GP or "GP's" or generalist*).ab. 72948
19 ((general or family or nurs*) adj1 (practice* or practitioner*)).tw,kf. 139716
20 NURSE PRACTITIONERS/ 18719
21 PRIMARY CARE NURSING/ 562
22 FAMILY NURSING/ 1561
23 HOME NURSING/ 8660
24 community health centers/ or substance abuse treatment centers/ or community mental health centers/15873
25 (homecare or home care or "care in the community").tw,kf. 24653
26 ((family or community or practi*) adj (medic* or doctor* or physician* or health* or nurs*)).tw,kf. 101566
27 ((in or at or based or own) adj2 (home or homes)).ab. 125585
28 Private Practice/ 8470
29 (private* adj1 practi*).tw. 12535
30 or/13-29 648401
31 12 and 30 611
32 meta-analysis/ or "systematic review"/ 293473
33 (systematic or structured or evidence or trials or studies).ti. and ((review or overview or look or examination or update* or summary).ti. or review.pt.) 304942
34 (0266-4623 or 1469-493X or 1366-5278 or 1530-440X or 2046-4053).is. 20689
35 meta-analysis.pt. or (meta-analys* or meta analys* or metaanalys* or meta synth* or meta-synth* or metasynth*).ti,ab,kf,hw. 286306
36 ((systematic or meta) adj2 (analys* or review)).ti,kf. or ((systematic* or quantitativ* or qualitativ* or methodologic*) adj5 (review* or overview*)).ti,ab,kf,sh. or ((quantitativ$ or qualitativ$) adj5 synth$).ti,ab,kf,hw. 402445
37 (integrative research review* or research integration).tw. or scoping review?.ti,kf. or (review.ti,kf,pt. and (trials as topic or studies as topic).hw.) or (evidence adj3 review*).ti,ab,kf. 233157
38 review.pt. and ((medline or medlars or embase or pubmed or scisearch or psychinfo or psycinfo or psychlit or psyclit or cinahl or electronic database* or bibliographic database* or computeri#ed database* or online database* or pooling or pooled or mantel haenszel or peto or dersimonian or der simonian or fixed effect or ((hand adj2 search*) or (manual* adj2 search*))).tw,hw. or (retraction of publication or retracted publication).pt.) 201561
39 (umbrella review or "review of reviews").tw. 1865
40 (rapid review or mixed method? review or scoping review or literature review).tw. 136898
41 or/32-40 832083

42 31 and 41 39
43 (PrEP or pre expos* prophyla* or preexpos* prophyla*).ti. 4686
44 41 and 43 224
45 42 or 44 241
46 afghanistan/ or exp africa/ or albania/ or andorra/ or antarctic regions/ or argentina/ or exp asia, central/ or exp asia, northern/ or exp asia, southeastern/ or exp atlantic islands/ or bahrain/ or bangladesh/ or Bhutan/ or bolivia/ or borneo/ or "bosnia and herzegovina"/ or brazil/ or bulgaria/ or exp central america/ or exp china/ or "commonwealth of independent states"/ or croatia/ or "democratic people's republic of korea"/ or ecuador/ or gibraltar/ or guyana/ or exp india/ or indonesia/ or iran/ or iraq/ or jordan/ or kosovo/ or kuwait/ or lebanon/ or liechtenstein/ or macau/ or "macedonia (republic)"/ or exp melanesia/ or moldova/ or monaco/ or mongolia/ or montenegro/ or nepal/ or netherlands antilles/ or new guinea/ or oman/ or pakistan/ or paraguay/ or peru/ or philippines/ or qatar/ or "republic of belarus"/ or romania/ or exp russia/ or saudi arabia/ or serbia/ or sri lanka/ or suriname/ or syria/ or taiwan/ or exp transcaucasia/ or ukraine/ or uruguay/ or united arab emirates/ or exp ussr/ or venezuela/ or yemen/ or sub-saharan africa*.ti,kf. 1297224
47 "organisation for economic co-operation and development"/496
48 australasia/ or exp australia/ or austria/ or exp baltic states/ or belgium/ or exp canada/ or chile/ or czech republic/ or colombia/ or europe/ or exp france/ or exp germany/ or greece/ or hungary/ or ireland/ or israel/ or exp italy/ or exp japan/ or korea/ or luxembourg/ or mexico/ or netherlands/ or new zealand/ or north america/ or poland/ or portugal/ or exp "republic of korea"/ or exp "scandinavian and nordic countries"/ or slovakia/ or slovenia/ or spain/ or switzerland/ or turkey/ or exp united kingdom/ or exp united states/3449325
49 european union/ 17445
50 developed countries/ 21253
51 or/47-50 3465007
52 46 not 51 1206134
53 45 not 52 215
54 Advisory Committees/10586
55 consensus/ 19633
56 Guideline/ or Practice Guideline/ 37395
57 health policy/ or international health regulations/ 71635
58 (advisory committee or advisory group or guideline or consensus statement).ti,ab,kf. 91029
59 (expert adj (review or recommendation)).mp. 2043
60 guideline.pt. 16548
61 or/54-60 216356
62 (NHS or NICE or "National Institute for Clinical Excellence" or "National Institute for Health and Care Excellence" or UK or GB or Great Britain or England or Ireland or Scotland or Wales).mp. 386369
63 61 and 62 10545
64 12 and 63 9
65 53 or 64 224
66 (infant* or child* or p?ediatr* or pregnan* or postpartum or post partum or postnatal* or post natal* or lactating).ti. 1531125
67 65 not 66 219
68 limit 67 to yr="2017 -Current" 162

**Supplementary Box S2 – Interview topic guide**

**Description of HIV testing**
Can you give me an overview of your HIV testing practices?
Who do you routinely offer HIV testing to/ how do you determine who to offer HIV tests to?
What symptoms or patient characteristics might make you order an HIV test?
Is there anything else that cues you to order an HIV test? E.g. information on computer screen / automatic prompts
What is the prevalence of HIV in the area covered by the practice? If not known, ask how they would find this out / who at the practice knows this information?
Do you have staff with a special interest in this area?
How much of a priority is HIV testing for your practice? If not a priority – why not?
Research shows that there have often been gaps in knowledge about guidelines. What do staff know about the BHIVA guidelines and government targets for ending HIV? What other resources and guidance do you use to guide your practice in this area? Is there scope for updating knowledge in this area?

**Barriers and facilitators to HIV testing**
We are interested in developing an intervention to increase testing and as such would like to know what works well in your practice, what doesn't work so well and what you feel could work better.
Thinking firstly about what would facilitate testing can you describe a situation in which it would be easy to offer and run an HIV test? [e.g. specialists]
Do you have any concerns about offering an HIV test?
If so, what?
Have you/the practice tried to improve testing before?
Research shows that barriers involve staff level, patient level and system level factors.
Commonly mentioned barriers involving staff include:

- Discomfort and lack of confidence approaching patients to offer an HIV test.
- Lack of knowledge and skills in sexual health counselling
- Insufficient knowledge of the clinical signs of HIV/how to identify high risk individuals and how to carry out a test.
- Not having sufficient time and resources to offer the test and follow up on the test.

Are these issues for your practice and have you been able to overcome any of them?
What other issues or barriers have you experienced?
Is there anything that might discourage or prevent you from offering and running an HIV test for a particular patient? (e.g. low perceived numbers of people living with HIV in the practice population, a feeling that the HIV test distresses patients (fear of offending), the consent process for testing).
Is there an issue now that patients can view test results on the NHS app?
Do you think some groups of people are more or less likely to be offered a test?
In terms of patient level factors frequently reported barriers include lack of culturally sensitive approaches, and patient concerns about confidentiality and the stigma they perceive around HIV preventing them from discussing HIV with their GPs (smaller rural practices may be more affected by issues of anonymity).
Are any of these barriers ones that you recognise may have had a negative impact on offering an HIV test by staff at your practice.
Has your practice found ways to overcome these barriers (through for example training or allocating testing to specific staff)?
Are there other barriers preventing your practice from increasing HIV testing.

**Approaches to increasing HIV testing**
Do you think there is a need to increase HIV testing in your practice?

What do you think could be done to increase HIV testing in your practice? (e.g. opt out as in maternity testing)
Are there difficulties in identifying individuals at risk?
The research shows that some approaches to increasing testing have systematised identifying people at risk of HIV (either using patient completed questionnaires or information held on electronic records). This would include identifying indicator conditions. Does your practice have a systematic way of identifying patients and if not would this sort of approach help? (use of a form has allowed providers to initiate conversations and provide a standard approach to enquiring about sexual health).
What type of support would you/ your practice need to increase HIV testing? [e.g. training etc]
What would this look like? How could this be introduced?
What training has been provided/is there an awareness of what is available? Are there issues with attending training (e.g. resources/time etc)
Would the type of test used support increased testing e.g. mouth swab versus blood test?
Could/should financial incentives be used to increase testing?
Is there a role for healthcare navigators or receptionists in this process [e.g. informing patients of practice policy for routine HIV testing]?
**Experiences of PrEP**
Can you tell me about any experience you have of discussing PrEP and how to access it with patients including referring patients to specialist sexual health services to access PrEP? If so, what prompts this [offering HIV test]?
**Barriers to accessing PrEP**
What might prevent you referring / supporting access to PrEP [lack of awareness and knowledge among clinical staff, unwillingness of staff to offer PrEP, unwillingness to discuss sexual practices, concern about risk compensation – riskier sexual practices, concerns about adherence to PrEP usage and management, lack of guidance or protocols]?
**Approaches to supporting access to PrEP**
What could be done to increase access to PrEP?
What role do you think general practice plays/should play in supporting access to PrEP? [signposting, advice, direct delivery from GP]
What role do you think general practices plays/should play in supporting PrEP maintenance care?
Which GP healthcare provider roles are most suited to supporting access to PrEP? [GPs, nurses, advanced nurse practitioners etc.]
Is there a role for healthcare navigators or receptionists in this process [e.g. informing patients about PrEP]? Could PrEP navigators be a useful role to support access?
What type of support would enable you and your practice to increase access to PrEP?
What would act as obstacles to engagement in these strategies?
What would support engagement in these strategies?
What would prevent general practice being more involved in supporting access to PrEP? [view PrEP delivery as role of sexual health clinics]

**Supplementary Table S3 – Behavioural analysis planning table**

| Barrier/ *facilitator* to target behaviour | Evidence for barrier/ facilitator/ intervention ingredient | Intervention ingredient | Behaviour change techniques / intervention function | Target construct (COM-B) |
|---|---|---|---|---|
| Target behaviour - Improving testing in GP practices in high-prevalence areas (testing all new patients and those undergoing blood tests for another reason). | | | | |
| Knowledge of guidelines and HIV prevalence is low among healthcare professions.<br><br>High prevalence practices appeared to lack processes or resources to test new patients or to test patients having a blood test for any other reason. | Interviews<br>Expert consensus<br>Literature:<br>Deblonde et al.2018<br>Tan and Black 2018<br>Serag et al. 2022<br>Mahendran et al. 2015<br><br>Interviews | Provide short and impactful information about the prevalence of HIV in the practice area, and the content of the guidelines for high/low prevalence areas. | Provide education / training | Psychological capability |
| Healthcare professionals lack resources to routinely test according to guidelines. | Interviews | Target high prevalence practices - offer test for new patients. Incentives may be required for any additional work undertaken other than work agreed through GP contract. | Incentivisation | Motivation |

| Barrier/ *facilitator* to target behaviour | Evidence for barrier/ facilitator/ intervention ingredient | Intervention ingredient | Behaviour change techniques / intervention function | Target construct (COM-B) |
|---|---|---|---|---|
| | | Simplify testing processes. | Incorporate HIV tests into existing tests | Physical opportunity |
| Healthcare professionals view routine testing as time consuming due to the belief that pre and post counselling is required.<br><br>There was insufficient time to discuss the test and address patient questions.<br><br>Lack of knowledge about rapid tests. | Interviews<br>Literature:<br>Deblonde et al. 2018<br>Davis et al. 2022<br>Loos et al. 2014<br><br>Interviews<br><br>Interviews | Interventions should persuade providers that lengthy pre and post counselling is not needed.<br><br>Simplifying the testing procedure | Provide education / training<br><br>Restructuring physical environment<br>Instruction on how to perform the behaviour | Psychological capability / physical opportunity<br><br>Physical opportunity / physical capability |
| Healthcare professionals lack confidence offering routine HIV tests due to a level of discomfort around discussing HIV with patients.<br><br>There was a lack of awareness about the stigma experienced by some minoritised groups in relation to HIV. | Interviews<br>Literature:<br>Deblonde et al. 2018<br>Tan and Black 2018<br>Desai et al. 2020<br>Pillay et al. 2024<br>Loos et al. 2014<br><br>Interviews<br>Literature<br>Deblonde et al. 2018 | Intervention should include skills training to help people to feel confident offering HIV test and to be able to answer questions and manage follow up. This should include training in culturally sensitive approaches. | Provide education / training<br>Ensure follow up protocol is known | Psychological capability / physical capability |

| Barrier/ *facilitator* to target behaviour | Evidence for barrier/ facilitator/ intervention ingredient | Intervention ingredient | Behaviour change techniques / intervention function | Target construct (COM-B) |
|---|---|---|---|---|
| Motivation to undertake routine tests is low due to perceived low prevalence of HIV and a social culture which focuses on treatment rather than prevention | Interviews | The intervention needs to persuade healthcare providers of the benefits of testing through providing information on the health benefits of early detection on prognosis and transmission of HIV including local data on missed opportunities to test among patients diagnosed late. Content should also provide information on "numbers needed to test" to ensure maintained motivation following repeated negative test results. Content may also highlight discrepancy between current and expected behaviour. | Provide education / training<br><br>Information about health consequences, information about social consequences, managing expectations<br><br>Provide feedback data on testing rates | Reflective motivation |

| Barrier/ *facilitator* to target behaviour | Evidence for barrier/ facilitator/ intervention ingredient | Intervention ingredient | Behaviour change techniques / intervention function | Target construct (COM-B) |
|---|---|---|---|---|
| Target behaviour - Improving testing in response to symptoms or indicator conditions in all areas | | | | |
| Knowledge of guidelines and prevalence is low among healthcare professionals. | Interviews<br><br>Literature:<br>Deblonde et al.2018<br>Tan and Black 2018<br>Serag et al. 2022<br>Mahendran et al. 2015 | Provide short and impactful information about the prevalence of HIV for the practice area, and the content of the guidelines for high/low prevalence areas. | Provide education / training | Psychological capability |
| Healthcare professionals may not recognise or be familiar with indicator conditions - and may not think about requesting an HIV test - particularly among those not considered typically high risk.<br><br>Some healthcare professionals thought pop-up reminders could be useful<br><br>Pop-up reminders would need to be designed well to overcome risk that it is closed down and ignored. | Interviews<br><br>Literature:<br>Desai et al. 2019<br>Davies et al. 2017<br>Bogers et al. 2021<br>Deblonde et al. 2018<br>Chadwick et al. 2021<br><br>Interviews<br><br>Interviews | Pop-up reminders must be co-designed to reduce the risk that they are ignored or switched off by healthcare professionals. | Prompts /cues | Physical capability |

| **Barrier/ *facilitator* to target behaviour** | **Evidence for barrier/ facilitator/ intervention ingredient** | **Intervention ingredient** | **Behaviour change techniques / intervention function** | **Target construct (COM-B)** |
|---|---|---|---|---|
| HIV tests currently not always included as standard when blood tests are requested and healthcare professionals do not always think about HIV when requesting blood tests. Wide variation in testing. | Interviews<br><br>Literature:<br>Deblonde et al. 2018 | Automatically include HIV testing within "packages" of tests<br><br>Use decision support system to prompt HIV testing | Environmental restructuring | Physical capability |
| Healthcare professionals do not (typically) have a systematic way of recording high-risk behaviours so may not always recognise opportunities for testing. | Interviews<br><br>Literature:<br>Tan and Black 2018<br>Chavez et al. 2018 | Intervention will not focus on identifying high risk behaviours | N/A | N/A |
| HIV testing is not always provided as part of STI testing.<br><br>Chlamydia and Gonorrhoea involve a different process to HIV testing. HIV testing may involve a further appointment with phlebotomist if the GP doesn't undertake it. | Interviews<br><br><br>Interviews | Include HIV within STI tests<br><br>Education about indicator conditions | Environmental restructuring | Physical capability |

| **Barrier/ *facilitator* to target behaviour** | **Evidence for barrier/ facilitator/ intervention ingredient** | **Intervention ingredient** | **Behaviour change techniques / intervention function** | **Target construct (COM-B)** |
|---|---|---|---|---|
| Healthcare professionals are reluctant to initiate conversations about HIV due to concerns about public/patient reactions | Interviews<br><br>Literature:<br>Tan and Black 2018<br>Fraser et al. 2022 | Combine practice level change with patient communication materials (text messages, posters, noticeboards) highlighting that HIV tests will be offered at all/any opportunity as standard.<br><br>To promote normalisation of testing add steps to existing processes e.g. add conversation about HIV testing/PrEP to sexual health consultations.<br>There is potential to add a pop-up for these consultations.<br><br>Provide leadership through practice champions. | Environmental restructuring | Social opportunity<br><br>Motivation |

| **Barrier/ *facilitator* to target behaviour** | **Evidence for barrier/ facilitator/ intervention ingredient** | **Intervention ingredient** | **Behaviour change techniques / intervention function** | **Target construct (COM-B)** |
|---|---|---|---|---|
| Target behaviour - healthcare professionals facilitating access to PrEP | | | | |
| Knowledge of PrEP is low among healthcare professionals | Interview<br><br>Literature:<br>Hillis et al. 2020<br>Conley et al. 2022<br>Hoornenborg et al. 2017<br>Pleuhs et al. 2020<br>Zhang et al. 2019<br>Matacotta et al. 2020<br>Mayer et al. 2020<br>Silapaswan et al. 2017<br>Wang et al. 2022<br>Zablotska et al. 2017<br>Bradley et al. 2019<br>Li et al. 2022<br>Siegler et al.2020 | Provide short and impactful information about PrEP | Provide education / training | Psychological capability |
| Most healthcare professionals do not discuss PrEP with patients due to lack of perceived opportunity and lack of knowledge about PrEP efficacy, safety and follow up - as well as viewing PrEP as something that should be delivered in sexual health service. | Interviews | Add steps to existing processes to discuss PrEP e.g. HIV test, STI screening, or long acting contraception consultations<br><br>Provide information about barriers patients can face in accessing sexual health services | Skills training<br>Add steps to existing processes | Psychological capability / physical opportunity |

| Barrier/ *facilitator* to target behaviour | Evidence for barrier/ facilitator/ intervention ingredient | Intervention ingredient | Behaviour change techniques / intervention function | Target construct (COM-B) |
|---|---|---|---|---|
| Perceived lack of time/resources to counsel patients, prescribe and carry out PrEP monitoring follow-up<br><br>Lack of knowledge results in discomfort in prescribing PrEP or limited awareness or understanding about guidelines. | Interview<br><br>Literature:<br>Pleuhs et al. 2020<br>Siegler et al. 2020<br>Silapaswan et al. 2017<br>Zhang et al. 2019<br>Li et al. 2022<br><br>Literature:<br>Cooper et al. 2021<br>Hillis et al. 2020<br>Hoornenborg et al. 2017Mayer et al. 2020<br>Allen et al. 2017<br>Saberi et al. 2022<br>Siegler et al. 2020<br>Silapaswan et al. 2017<br>Yusuf et al. 2020 | | | |
| Most healthcare professionals have not signposted patients to specialist services for PrEP due to lack of perceived opportunity, lack of knowledge and assuming people who need it already know about it | Interviews | Implement communication systems to signpost patients to specialist services using information in GP practices, text messages and links to information on websites about PrEP (following | Environmental restructuring | Physical/Social opportunity |

| Barrier/ *facilitator* to target behaviour | Evidence for barrier/ facilitator/ intervention ingredient | Intervention ingredient | Behaviour change techniques / intervention function | Target construct (COM-B) |
|---|---|---|---|---|
| | | consultations or as part of a campaign) | | |
| Healthcare professionals have mistaken beliefs about who is at risk. | Literature:<br>Matacotta et al. 2020<br>Hoornenborg et al. 2017 | | Provide education / training | Psychological capability |
| Healthcare professionals are concerned about behavioural and health consequences – belief that PrEP will lead to risky behaviour. | Pleuhs et al. 2020<br>Zhang et al. 2019<br>Silapaswan et al. 2017<br>Li et al. 2022<br>Cooper et al. 2021<br>Turner et al. 2018 | | | |
| Discomfort about discussing sexual activities/history with patients | Pleuhs et al. 2020<br>Matacotta et al. 2020 | | | |
| Negative influence of healthcare professionals race, gender and age biases – interpersonal stigma). | Pleuhs et al. 2020 | | | |

**Supplementary Figure S4 – Logic Model**

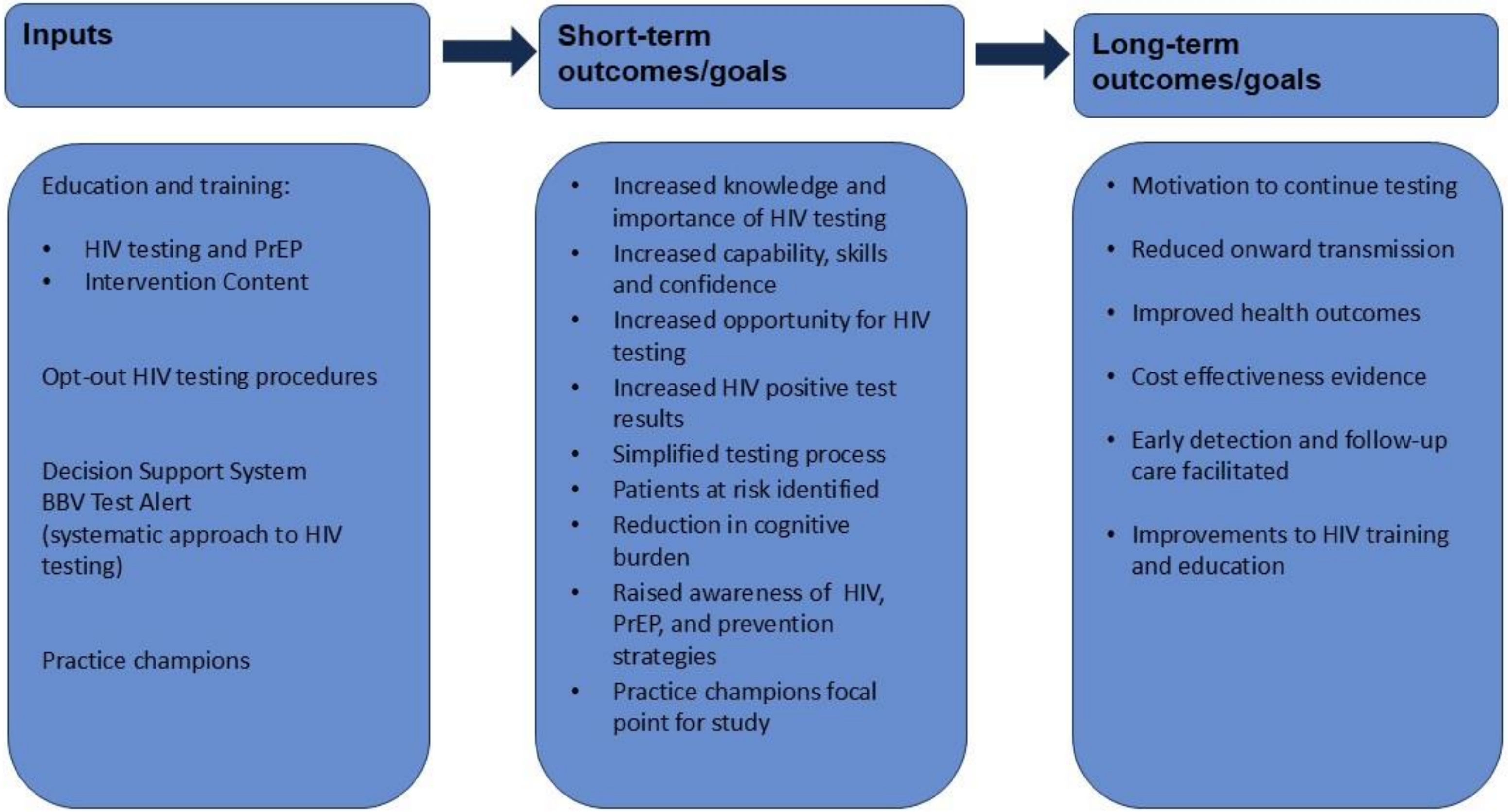

**Supplementary Table S5 – Guiding Principles**

| Context-specific behavioural issues | Design objective | Intervention feature |
|---|---|---|
| Motivation to offer HIV tests may be low due to multiple competing demands and a need to prioritise all/alternative disease/ conditions. Awareness of HIV prevalence and testing guidelines may reduce perceived importance (given low numbers of positive results). | To motivate healthcare professionals to offer HIV testing to people in response to guideline recommendations. | Highlight the need for HIV testing through informing healthcare professionals about the local prevalence and national guideline recommendations<br><br>Highlight the importance of early detection and treatment including UNAIDS elimination goals and Undetectable=Untransmissable message. |
| Healthcare professionals have limited time and resources to receive and deliver additional interventions. Given the historical context, many have expectations that offering an HIV test will be overly time-consuming. | To provide easily accessible intervention components that are suitable for a range of staff groups | Offer brief online training that healthcare professionals can view at a time that suits them<br><br>Emphasise that tests can be offered and delivered quickly and easily without the need for lengthy counselling.<br><br>Create practice champion role to support introduction of initiatives to support HIV testing and discussions about PrEP. |
| Healthcare professionals have low self-efficacy to deliver HIV testing and have discussions about PrEP in general practice. | To empower and support acceptable and feasible implementation of HIV testing and discussions about PrEP in general practice | Provide patient electronic health record (EHR) system prompt to facilitate HIV testing in response to system based prompts when patients present with indicator conditions or symptoms.<br><br>Use patient information to normalise HIV testing and raise awareness of PrEP to reassure healthcare professionals that HIV testing is acceptable to patient populations and will not result in length consultations.<br><br>Automate addition of HIV testing. |

| Context-specific behavioural issues | Design objective | Intervention feature |
|---|---|---|
| Healthcare professionals perceive specialist sexual health services as more appropriate settings for HIV testing and delivery of PrEP. | To motivate healthcare professionals to view HIV testing and discussions about PrEP as part of their role in general practice. | Highlight data on the number of times patients diagnosed late are seen in general practice.<br>Highlight barriers to accessing sexual health services experienced by patients resulting in inequitable access to HIV testing and PrEP and the central role GPs can play. |

**Supplementary Table S6 – Developing the intervention**

| COM-B | Barriers | Facilitators |
|---|---|---|
| Capability | • Lack of knowledge, awareness, skills to test for HIV or support access to PrEP<br>• Lack of systematic approaches to HIV testing and discussion of PrEP | • Education<br>• Embed HIV testing in existing systems<br>• Use electronic health record prompt |
| Opportunity | • Time is perceived as a barrier to carrying out HIV testing<br>• Perceptions about lengthy consent<br>• Shortages of blood appointments/staff to carry out blood tests<br>• Time is a barrier to discussing PrEP | • Simplify testing processes (opt-out)<br>• Educate staff that lengthy consent is not required<br>• Incorporate HIV tests into existing blood tests automatically<br>• Incorporate discussions about PrEP into existing processes e.g. STI screening or long-acting contraception consultations |
| Motivation | • Reluctance to initiate discussions about HIV tests and PrEP | • Normalise processes, reduce stigma among staff and patients |

| COM-B | Barriers | Facilitators |
| --- | --- | --- |
| | • Stigma | • Provide leadership through practice champions<br>• Persuade healthcare professionals of the benefits of testing and signposting to PrEP<br>• Consider incentives |
